# Selection of optimum formulation of RBD-based protein sub-unit covid19 vaccine (Corbevax) based on safety and immunogenicity in an open-label, randomized Phase-1 and 2 clinical studies

**DOI:** 10.1101/2022.03.08.22271822

**Authors:** Subhash Thuluva, Vikram Paradkar, Kishore Turaga, SubbaReddy Gunneri, Vijay Yerroju, Rammohan Mogulla, Mahesh Kyasani, Senthil Kumar Manoharan, Guruprasad Medigeshi, Janmejay Singh, Heena Shaman, Chandramani Singh, A Venkateshwar Rao

## Abstract

**Background:** We present the data from an open-label study involved in the selection of optimum formulation of RBD-based protein sub-unit COVID-19 vaccine.

**Methods:** The randomized Phase-1/2 trial followed by a Phase-2 trial was carried out to assess safety and immunogenicity of different formulation of COVID-19 vaccine (Corbevax) and select an optimum formulation for a phase 3 study. Healthy adults without a history of Covid-19 vaccination or SARS-CoV-2 infection, were enrolled.

**Findings:** Low incidence of adverse events were reported post-vaccination of different Corbevax formulations and majority were mild in nature and no Grade-3 or serious adverse events were observed. All formulations in Phase-1/2 study showed similar profile of humoral and cellular immune-response with higher response associated with increasing CpG1018 adjuvant content at same RBD protein content. Hence, high concentration of CpG1018 was tested in phase-2 study, which showed significant improvement in immune-responses in terms of anti-RBD-IgG concentrations, anti-RBD-IgG1 titers, nAb-titers and cellular immune-responses while maintaining the safety profile. Interestingly, binding and neutralizing antibody titers were persisted consistently till 6 months post second vaccine dose.

**Interpretations:** Corbevax was well tolerated with no observed safety concerns. Neutralizing antibody titers were suggestive of high vaccine effectiveness compared with human convalescent plasma or protective thresholds observed during vaccine efficacy trials of other COVID-19 vaccines. The study was prospectively registered with clinical trial registry of India-CTRI/2021/06/034014 and CTRI/2020/11/029032.

**Funding:** Bill & Melinda Gates Foundation, BIRAC-division of Department of Biotechnology, Govt of India, and the Coalition for Epidemic Preparedness Innovations funded the study.

## INTRODUCTION

Coronavirus disease (Covid-19) is caused by the Severe Acute Respiratory Syndrome Coronavirus 2 (SARS-CoV-2)^1^. Multiple vaccines have either been developed or are under development to prevent infection and reduce disease severity. Majority of vaccines utilize the Spike protein or entire virus (inactivated virus vaccines) as the source of antigen.

Protein sub-unit vaccines consist of either whole protein or specific regions of the protein from the pathogen containing key B- and T-cell epitopes combined with adjuvant^2^. Multiple research groups have shown that Receptor Binding Domain (RBD) of the spike protein can generate excellent immune-response in terms of nAb-titers against SARS-COV-2 virus^3–6^. Biological E developed RBD-based sub-unit vaccine with Aluminum Hydroxide and CpG1018 as adjuvants. Baylor College of Medicine/Texas Children’s Hospital produced and licensed the recombinant *Pichia Pastoris* strain expressing RBD protein to Biological E^7–11^ and Dynavax Inc. supplied the CpG1018 adjuvant. After satisfactory results from pre-clinical and development studies, clinical studies were planned to finalize the vaccine formulation for late-stage clinical studies.

The study describes the clinical studies that led to the optimum formulation of the vaccine (Corbevax). The Phase-1/2 study was conducted to assess the key role played by RBD and the adjuvant CpG1018. The subsequent Phase-2 study was conducted to confirm the safety and immunogenicity of the optimum formulation. In addition, persistence of immune-response till 6-months post completion of primary-vaccination series is also presented for the formulations used in the Phase-1/2 study.

## METHODS

### Study Population and Study Design

Overall, 1497 subjects were screened as part of two studies and 460 subjects (n=360 in Phase-1/2 and n=100 in Phase-2 study) were vaccinated with various formulations of Corbevax vaccine (Table 1). Subject disposition in phase-1/2 and phase-2 study are illustrated in Figure-1 and 2 respectively.

**Figure 1:**
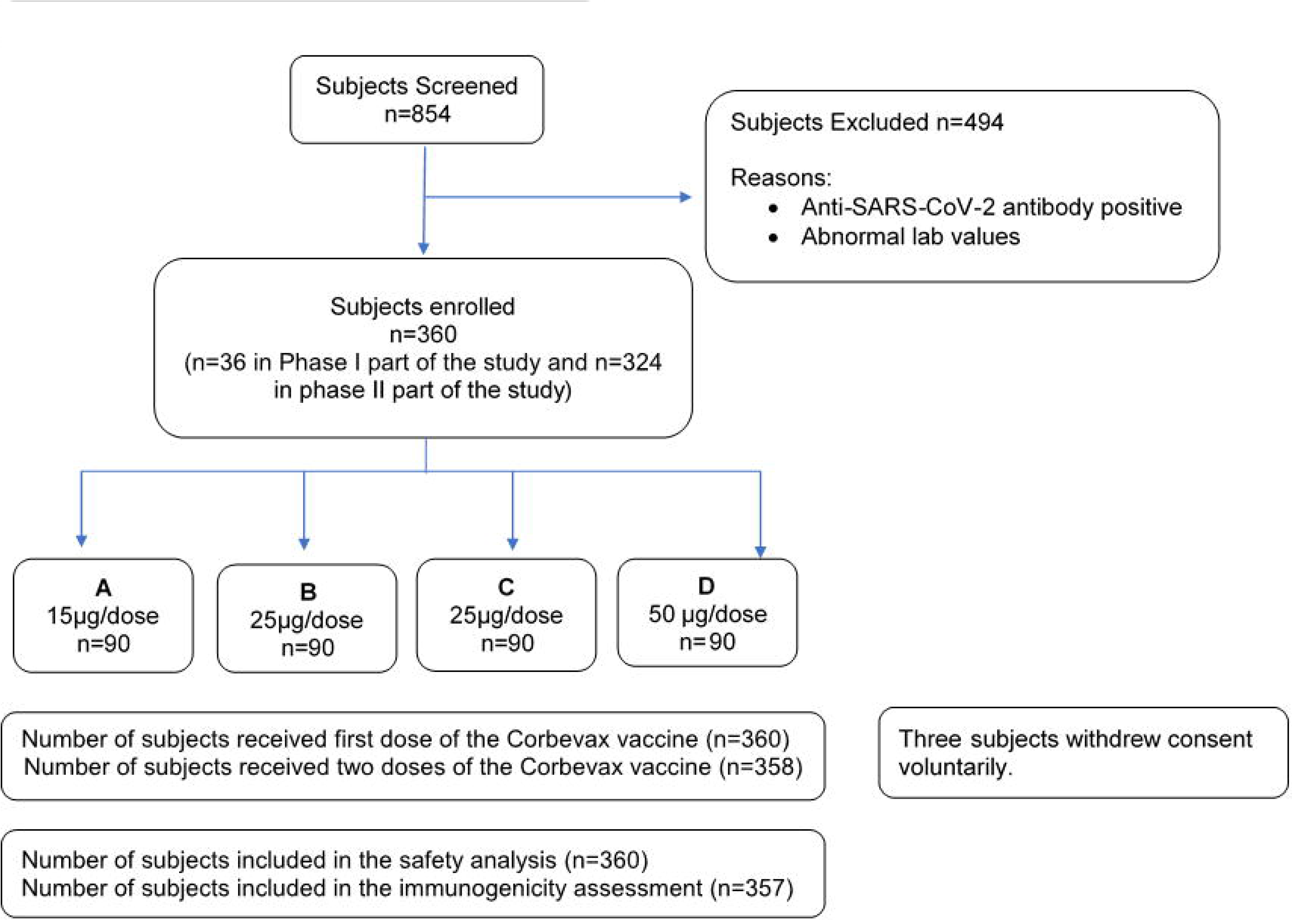
Subject disposition in Phase 1/2 study

**Figure 2:**
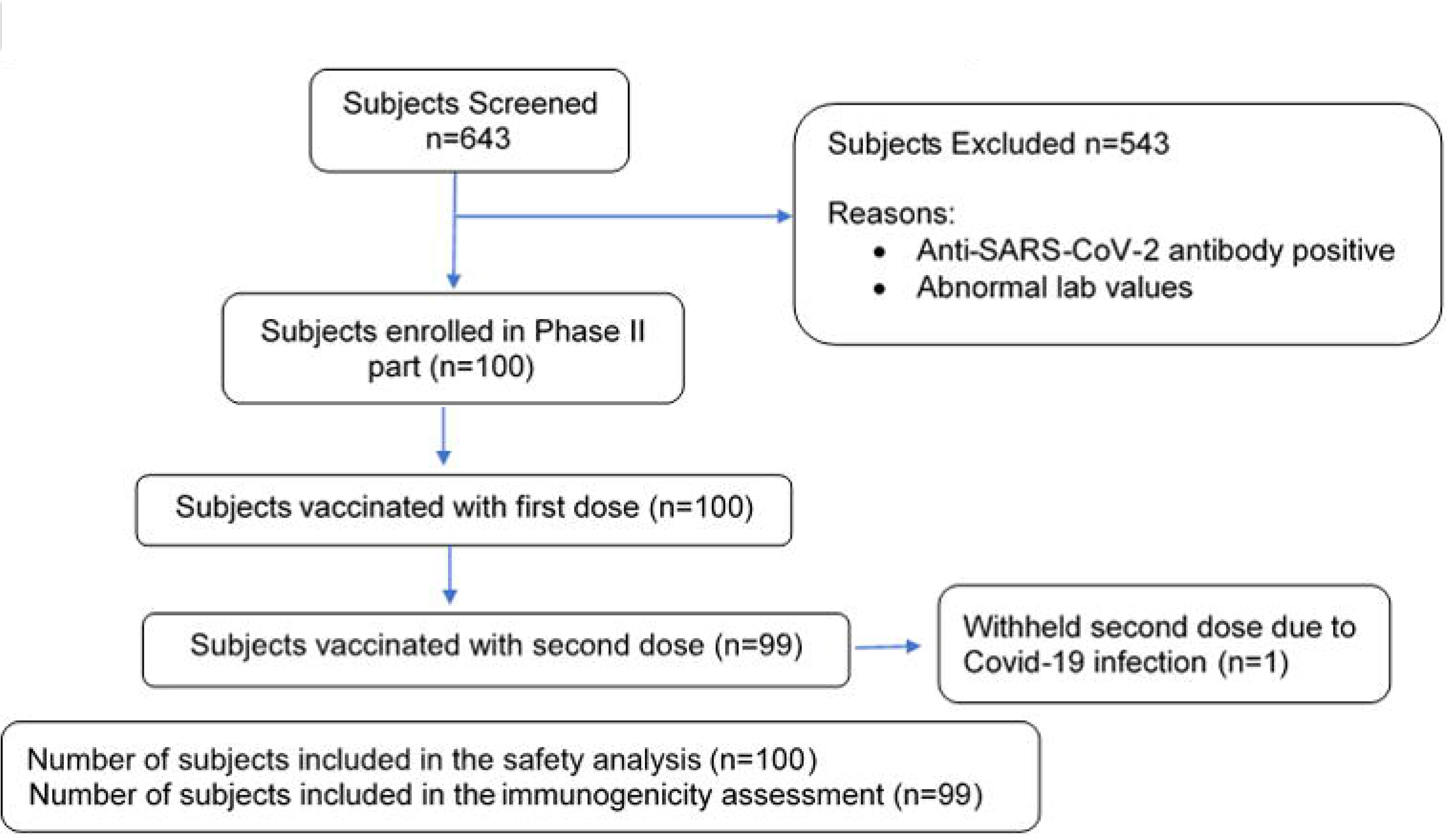
Subject disposition in Phase II part of Phase II/III study

**Table 1:** Composition of the different formulations of Corbevax

| Component details | Phase-1/ 2 study |  |  |  | Phase-2 study |
| --- | --- | --- | --- | --- | --- |
|  | A | B | C | D | E |
| RBD antigen of SARS-CoV-2 (Covid-19) | 50 µg | 25 µg | 25 µg | 15 µg | 25 µg |
| Aluminium Hydroxide gel as Al<sup>+++</sup> | 750 µg | 750 µg | 500 µg | 750 µg | 750 µg |
| CpG 1018 | 500 µg | 500 µg | 250 µg | 500 µg | 750 µg |
| Buffer (Tris and NaCl in WFI) | q.s to 0.5 mL | q.s to 0.5 mL | q.s to 0.5 mL | q.s to 0.5 mL | q.s to 0.5 mL |

#### Study Design

The Phase-1/2 and Phase-2 studies were carried out in 5 and 7 centers across India respectively between 11-November-2020 to 20-August-2021. Studies are prospective, open-label, randomized (phase-1/2) to assess best vaccine formulation based on safety, tolerability, reactogenicity and immunogenicity in Covid-19 RT-PCR and sero-negative subjects.

Phase-1/2 study had 360 healthy volunteers were randomly assigned into four groups to receive 0.5 mL dose of 4 different Corbevax formulations designated as A, B, C & D in a 2-dose schedule with 28 days’ interval between doses (Figure-1). All the subjects are under follow-up till one year after the second-dose. Six-month follow-up data also included in this manuscript. Data from Phase-1/2 study indicated significant potential to enhance overall immune-response by increasing CpG1018 adjuvant. After completing the formulation development work, the optimum Corbevax formulation consisting of RBD antigen (25µg) + Aluminium Hydroxide (750µg) + CpG 1018 (750µg) per human dose of 0.5mL was tested in Phase-2 study (formulation-E, Table 1), a total of 100 healthy subjects were enrolled. All participants will be followed up for 12-months after second-dose of vaccine administration.

Eligible participants were healthy men and women aged 18-65 years at the time of 1^st^ vaccination for Phase-1/2 and Phase-2 studies. They had to be negative to SARS-CoV-2 infection and anti-SARS-CoV-2 antibody prior to enrolment. (detailed eligibility criteria are described in supplementary material).

The Investigational Review Board or Ethics Committee at each study site approved the protocol. Details of all Ethics Committees / Institutional Review Boards were listed as supplementary information. The study was conducted in accordance with the principles defined in the Declaration of Helsinki, International Conference on Harmonization and the local regulatory guidelines. Written informed consents were obtained from all healthy volunteers prior to the enrollment.

#### Randomization and masking

Equal randomization of subjects into different formulation groups was performed using Interactive Web Response System which containing randomization number and intended allocation. The allotment of randomization number is initiated by assigning first randomization no. e.g.: EA001 (E-enrollment; A-site code; 001-number of enrolled subject) and this continued in the same serial order till all the subjects are randomized. Masking is not applicable to open label studies.

#### Safety assessments

Each subject was under direct observation for any immediate local and systemic adverse reactions up to 120-minutes of post-vaccination by the investigator. All subjects were provided with subject diary and trained on how to observe and capture adverse symptoms post-vaccination for next seven consecutive calendar days. Only the principal investigator or co-investigator performed assessment of causality of reported symptoms.

#### Immunological assays

SARS-CoV-2 antibody concentrations for diagnostic purpose were measured using Diasorin kit^12^. Anti-RBD antibody responses and IgG-subclass responses were measured using validated ELISA-method. SARS-COV-2 neutralizing antibody titers were measured using MicroNeutralizationAssay (MNA) and PseudovirusNeutralizationAssay (PNA) methods and cellular immune-responses were assessed by cytokine secretion using TrueCulture® tubes coated with SARS-COV-2 peptides. Details on assays and methods used for assessing immunological parameters were descried as supplementary information.

### Statistical Methods

#### Statistical Analyses

All data are presented using descriptive statistics. Demographic and primary safety analyses were based on total vaccinated population. Full-analysis-set (FAS) included subjects who provided informed consent for participation. Intent-to-treat (ITT) analysis set included all subjects from FAS who had received both the doses of study vaccine. Per protocol (PP) analysis set included all subjects from ITT set without any major protocol deviations. All subjects entered into the study and who received at least one dose of study vaccine were included in the safety-analysis. All immunological data was log-transformed to obtain a log-normal distribution. All the statistical analyses were conducted using SAS^®^ 9.4 or higher (SAS Institute, Cary NC).

## RESULTS

### Study Population

Subjects’ demographics and baseline characteristics are described in Table 2 and subjects’ disposition is shown in Figure-1 and 2.

**Table 2.** Demographics and Baseline Characteristics of participants in Phase-1/ 2 study and Phase-2 of Phase-2/3 Study

| Parameter/<br>Statistics/Category | Phase-1/2 study |  |  |  | Overall (N=360) | Phase-2 Study<br>E<br>(n=100) |
| --- | --- | --- | --- | --- | --- | --- |
|  | A<br>(N=90) | B<br>(N=90) | C<br>(N=90) | D<br>(N=90) |  |  |
| Age (Year) |  |  |  |  |  |  |
| Mean ±SD | 35.0±8.7 | 35.0±9.1 | 32.7±8.1 | 34.7±6.7 | 34.4±8.2 | 33.2 ±8.41 |
| Range (Min:max) | (20.0:63.0) | (19.0:60.0) | (18.0:53.0) | (20.0:50.0) | (18.0:63.0) | 18.0; 52.0 |
| Gender, N<sub>1</sub> (%) |  |  |  |  |  |  |
| Male | 74 (82.22%) | 75 (83.33%) | 74 (82.22%) | 82 (91.11%) | 305 (84.72%) | 86 (86.00%) |
| Female | 16 (17.78%) | 15 (16.67%) | 16 (17.78%) | 8 (8.89%) | 55 (15.28%) | 14 (14.00%) |
| Height (Cms) |  |  |  |  |  |  |
| Mean ±SD | 166.8±8.9 | 166.9±7.8 | 165.7±8.4 | 165.8±6.8 | 166.3±8.0 | 167.7 ±7.23 |
| Range (Min:max) | (146.0:186.0) | (147.0:185.0) | (135.0:182.0) | (152.0:179.6) | (135.0:186.0) | 148.0; 182.0 |
| Weight (Kgs) |  |  |  |  |  |  |
| Mean ±SD | 69.8±10.0 | 68.8±11.6 | 67.5±11.2 | 68.9±9.7 | 68.8±10.7 | 65.7 ±10.90 |
| Range (Min:max) | (48.0:88.7) | (35.0:105.0) | (35.0:89.4) | (50.0:92.0) | (35.0:105.0) | 40.0; 96.0 |
| BMI (Kg/m<sup>2</sup>) |  |  |  |  |  |  |
| Mean ±SD | 25.1±2.6 | 24.6±3.2 | 24.5±3.2 | 25.1±3.1 | 24.8±3.0 | NR |
| Range (Min:max) | (18.3:29.2) | (15.0:30.7) | (18.3:30.9) | (17.1:33.6) | (15.0:33.6) |  |

**Note:** Percentages were calculated using column header group count as denominator.
The Age were calculated using following formula: Age (at vaccination) =round of ((Vaccination Date-Birth Date)/365.25).
BMI= Weight/(Height (in mts)<sup>2</sup>), N<sub>1</sub>: Subject Count, N: Sample Size.

### Safety Findings

In Phase-1/2 study, AEs were reported in 42 subjects (11.67%), in the range of 8.89-15.56% of subjects in different formulations groups. Least number of subjects with AEs reported in “B” formulation and highest was reported in “D” formulation (Table 3) In the Phase-2 study, a total of 27 (27.00%) subjects reported AEs post-vaccination. No AEs were reported in any subject within 120-minutes’ post-vaccination. None of the AEs were serious, or of Grade-3 severity. Details of solicited local and systemic AEs and unsolicited AEs were listed in Table-3 (Phase-1/2) and Table-5 (Phase-2). All the reported AEs were mild to moderate in severity and most of the events were considered related to the study vaccine. MAAEs were reported in 10 subjects (2.78%) and 6 subjects (6%) in Phase-1/2 and phase-2 studies respectively, of which none were reported as serious (Table 4).

**Table 3:** Adverse events and reactions in the Phase-1/2 study

| **N<sub>1</sub> (%) [95% CI] n** | **A**<br>**(N=90)** | **B**<br>**(N=90)** | **C**<br>**(N=90)** | **D**<br>**(N=90)** | **Overall**<br>**(N= 360)** |
| --- | --- | --- | --- | --- | --- |
| **Overall Adverse Events** |  |  |  |  |  |
| **Any AE** | 11 (12.22%)<br>[6.26, 20.82] 11 | 9 (10.00%)<br>[4.68, 18.14] 9 | 8 (8.89%)<br>[3.92, 16.77] 10 | 14 (15.56%)<br>[8.77, 24.72] 16 | 42 (11.67%)<br>[8.54, 15.44] 46 |
| **Any Local AE** | 7 (7.78%)<br>[3.18, 15.37] 7 | 3 (3.33%)<br>[0.69, 9.43] 3 | 3 (3.33%)<br>[0.69, 9.43] 4 | 12 (13.33%)<br>[7.08, 22.13] 12 | 25 (6.94%)<br>[4.54, 10.08] 26 |
| **Solicited Local Adverse Events** |  |  |  |  |  |
| Injection site erythema | 1 (1.11%)<br>[0.03, 6.04] 1 | 0 (0.00%) [NE] 0 | 0 (0.00%) [NE] 0 | 0 (0.00%) [NE] 0 | 1 (0.28%)<br>[0.01, 1.54] 1 |
| Injection site pain | 6 (6.67%)<br>[2.49, 13.95] 6 | 3 (3.33%)<br>[0.69, 9.43] 3 | 2 (2.22%)<br>[0.27, 7.80] 3 | 10 (11.11%)<br>[5.46, 19.49] 10 | 21 (5.83%)<br>[3.65, 8.78] 22 |
| Injection site swelling | 0 (0.00%) [NE] 0 | 0 (0.00%) [NE] 0 | 1 (1.11%)<br>[0.03, 6.04] 1 | 2 (2.22%)<br>[0.27, 7.80] 2 | 3 (0.83%)<br>[0.17, 2.42] 3 |
| **Any Systemic AEs** | 4 (4.44%)<br>[1.22, 10.99] 4 | 5 (5.56%)<br>[1.83, 12.49] 5 | 5 (5.56%)<br>[1.83, 12.49] 6 | 4 (4.44%)<br>[1.22, 10.99] 4 | 18 (5.00%)<br>[2.99, 7.79] 19 |
| **Solicited Systemic Adverse Events** |  |  |  |  |  |
| Chills | 0 (0.00%) [NE] 0 | 0 (0.00%) [NE] 0 | 1 (1.11%)<br>[0.03, 6.04] 1 | 0 (0.00%) [NE] 0 | 1 (0.28%)<br>[0.01, 1.54] 1 |
| Pyrexia | 3 (3.33%)<br>[0.69, 9.43] 3 | 2 (2.22%)<br>[0.27, 7.80] 2 | 2 (2.22%)<br>[0.27, 7.80] 2 | 1 (1.11%)<br>[0.03, 6.04] 1 | 8 (2.22%)<br>[0.96, 4.33] 8 |
| Myalgia | 0 (0.00%) [NE] 0 | 1 (1.11%)<br>[0.03, 6.04] 1 | 1 (1.11%)<br>[0.03, 6.04] 1 | 0 (0.00%) [NE] 0 | 2 (0.56%)<br>[0.07, 1.99] 2 |
| Headache | 1 (1.11%)<br>[0.03, 6.04] 1 | 2 (2.22%)<br>[0.27, 7.80] 2 | 0 (0.00%) [NE] 0 | 3 (3.33%)<br>[0.69, 9.43] 3 | 6 (1.67%)<br>[0.61, 3.59] 6 |
| Urticaria | 0 (0.00%) [NE] 0 | 0 (0.00%) [NE] 0 | 2 (2.22%)<br>[0.27, 7.80] 2 | 0 (0.00%) [NE] 0 | 2 (0.56%)<br>[0.07, 1.99] 2 |
| **Any Unsolicited Systemic AE** |  |  |  |  |  |
| Dyspepsia | 0 (0.00%) [NE] 0 | 1 (1.11%)<br>[0.03, 6.04] 1 | 0 (0.00%) [NE] 0 | 0 (0.00%) [NE] 0 | 1 (0.28%)<br>[0.01, 1.54] 1 |

**Table 4:** Medically attended AEs by SOC and PT in TVG

| **Phase-1/2** |  |  |  |  |  |
| --- | --- | --- | --- | --- | --- |
| **SOC/PT**<br>**N<sub>1</sub> (%) [95% CI] n** | **Treatment Groups** |  |  |  | **OVERALL**<br>**(N=360)** |
|  | **A**<br>**(N=90)** | **B**<br>**(N=90)** | **C**<br>**(N=90)** | **D**<br>**(N=90)** |  |
| Gastrointestinal disorders | 0 (0.00%) [NE] 0 | 1 (1.11%)<br>[0.03, 6.04] 1 | 0 (0.00%) [NE] 0 | 0 (0.00%) [NE] 0 | 1 (0.28%)<br>[0.01, 1.54] 1 |
| Dyspepsia | 0 (0.00%) [NE] 0 | 1 (1.11%)<br>[0.03, 6.04] 1 | 0 (0.00%) [NE] 0 | 0 (0.00%) [NE] 0 | 1 (0.28%)<br>[0.01, 1.54] 1 |
| General disorders and<br>administration site conditions | 1 (1.11%)<br>[0.03, 6.04] 1 | 1 (1.11%)<br>[0.03, 6.04] 1 | 1 (1.11%)<br>[0.03, 6.04] 1 | 1 (1.11%)<br>[0.03, 6.04] 1 | 4 (1.11%)<br>[0.30, 2.82] 4 |
| Pyrexia | 1 (1.11%)<br>[0.03, 6.04] 1 | 1 (1.11%)<br>[0.03, 6.04] 1 | 1 (1.11%)<br>[0.03, 6.04] 1 | 1 (1.11%)<br>[0.03, 6.04] 1 | 4 (1.11%)<br>[0.30, 2.82] 4 |
| Nervous system disorders | 1 (1.11%)<br>[0.03, 6.04] 1 | 0 (0.00%) [NE] 0 | 0 (0.00%) [NE] 0 | 2 (2.22%)<br>[0.27, 7.80] 2 | 3 (0.83%)<br>[0.17, 2.42] 3 |
| Headache | 1 (1.11%)<br>[0.03, 6.04] 1 | 0 (0.00%) [NE] 0 | 0 (0.00%) [NE] 0 | 2 (2.22%)<br>[0.27, 7.80] 2 | 3 (0.83%)<br>[0.17, 2.42] 3 |
| Skin and subcutaneous tissue<br>disorders | 0 (0.00%) [NE] 0 | 0 (0.00%) [NE] 0 | 2 (2.22%)<br>[0.27, 7.80] 2 | 0 (0.00%) [NE] 0 | 2 (0.56%)<br>[0.07, 1.99] 2 |
| Urticaria | 0 (0.00%) [NE] 0 | 0 (0.00%) [NE] 0 | 2 (2.22%)<br>[0.27, 7.80] 2 | 0 (0.00%) [NE] 0 | 2 (0.56%)<br>[0.07, 1.99] 2 |
| **Phase-2** |  |  |  |  |  |
| **System Organ Class**<br>**Preferred Term** | **E**<br>**(N=100)**<br>**N1(%), 95% CI, n** |  |  |  |  |
| Subjects with medically attended adverse events | 6 (6.0) [0.02, 0.13] 12 |  |  |  |  |
| Infections and infestations | 3 (3.0) [0.01, 0.09] 3 |  |  |  |  |
| COVID-19 | 1 (1.0) [0.00, 0.05] 1 |  |  |  |  |
| Nasopharyngitis | 1 (1.0) [0.00, 0.05] 1 |  |  |  |  |
| Pharyngitis | 1 (1.0) [0.00, 0.05] 1 |  |  |  |  |
| General disorders and administration site conditions | 2 (2.0) [0.00, 0.07] 4 |  |  |  |  |
| Pain | 2 (2.0) [0.00, 0.07] 2 |  |  |  |  |

| System Organ Class<br>Preferred Term | E<br>(N=100)<br>N1(%), 95% CI, n |
| --- | --- |
| Pyrexia | 2 (2.0) [0.00, 0.07] 2 |
| Musculoskeletal and connective tissue disorders | 1 (1.0) [0.00, 0.05] 1 |
| Pain in extremity | 1 (1.0) [0.00, 0.05] 1 |
| Nervous system disorders | 1 (1.0) [0.00, 0.05] 1 |
| Lethargy | 1 (1.0) [0.00, 0.05] 1 |

Note: Percentages were calculated using column header count as denominator.
N1: Subject count in specified category. N= Total number of subject. n= Event Count (One subject may be counted more than once),
NE: Not Estimable. 95% CI was based on percentage calculated by Clopper-Pearson Method.

**Table 5:** Adverse events and reactions in the Phase-2 study

| **N<sub>1</sub> (%) [95% CI] n** | **Corbevax**<br>**(N= 100)** |
| --- | --- |
| **Overall adverse events** |  |
| Any Adverse events | 27 (27.0) [0.19, 0.37] 105 |
| Solicited adverse event | 20 (20.0) [0.13, 0.29] 52 |
| Unsolicited adverse events | 10 (10.0) [0.05, 0.18] 17 |
| Medically Attended Adverse Events | 6 (6.0) [0.02, 0.13] 12 |
| Adverse Events Following Immunization | 24 (24.0) [0.16, 0.34] 47 |
| **Local Adverse events** |  |
| Injection site pain | 14 (14.0) [0.08, 0.22] 39 |
| Injection site erythema | 2 (2.0) [0.00, 0.07] 5 |
| Injection site swelling | 1 (1.0) [0.00, 0.05] 2 |
| **Systemic Adverse events** |  |
| Pyrexia | 9 (9.0) [0.04, 0.16] 15 |
| Fatigue | 4 (4.0) [0.01, 0.10] 9 |
| Pain | 2 (2.0) [0.00, 0.07] 2 |
| Gastrointestinal disorders | 4 (4.0) [0.01, 0.10] 6 |
| Musculoskeletal and connective tissue disorders | 4 (4.0) [0.01, 0.10] 10 |
| Nervous system disorders | 4 (4.0) [0.01, 0.10] 13 |
| Infections and infestations | 3 (3.0) [0.01, 0.09] 3 |
| Reproductive system and breast disorders | 1 (1.0) [0.00, 0.05] 1 |
| Respiratory, thoracic and mediastinal disorders | 1 (1.0) [0.00, 0.05] 3 |
| N1: Subject count in specified category; N= Total number of subject; n= Event Count; CI: confidence interval. |  |

No abnormal laboratory values, vital signs or physical examination were reported as clinically significant. There was no SAE reported during the reported study period.

### Immunogenicity Findings

#### Phase-1/2 study

Data is presented for the four subject cohorts that received the four different vaccine formulations, designated as A, B, C and D respectively. RBD-IgG The anti-RBD-IgG concentrations increased moderately after the first dose and then significantly after the second-dose and the values plateaued between Day42 and 56 (Figure-3). Percent seroconversion was highest (90%) for “B” formulation (Table-6). IgG1 (Th1-skewed) and IgG4 (Th2-skewed) titers were also measured from all four formulation cohorts. IgG1 titers were significant increased in all four cohorts with the highest GMT (2940 at day56) and GMFR (39.7) observed for B-formulation. The Day0 titers were very low for both isotypes in all the cohorts. In comparison, minor increase was observed in IgG4 titers for the four cohorts at day56 (Table 7)nAb-titers. Nexelis had conducted testing of 273 convalescent plasma samples collected from RT-PCR positive COVID-19 subjects with a range of disease severity, from asymptomatic to hospitalized patients. The GMT via PNA method for these convalescent sera panel was 126 and is included in the Figure-4 at day0,28,42 and day56. The nAb-titer trend is similar to the anti-RBD-IgG concentrations at corresponding time-points with highest NT50 titers, 130 at day42 induced by formulation-B. Figure-5 shows the GMT’s for the nAb-titers measured via MNA for the four cohorts at Day0 and Day56 time-points which shows similar increase in the overall titer; however, GMFR and % seroconversion were not calculated as the Day0 titers were measured for only a subset of randomly selected subjects by the MNA method. UKHSA^13^ conducted testing of 32 convalescent plasma samples collected from RT-PCR positive COVID19 subjects with severe disease and the GMT via MNA method for these convalescent sera panel was 522.. Formulation-B induced high NT50 (60 at day0 Vs 537 at day56) compared to other formulations tested (Figure-5).

**Figure 3:**
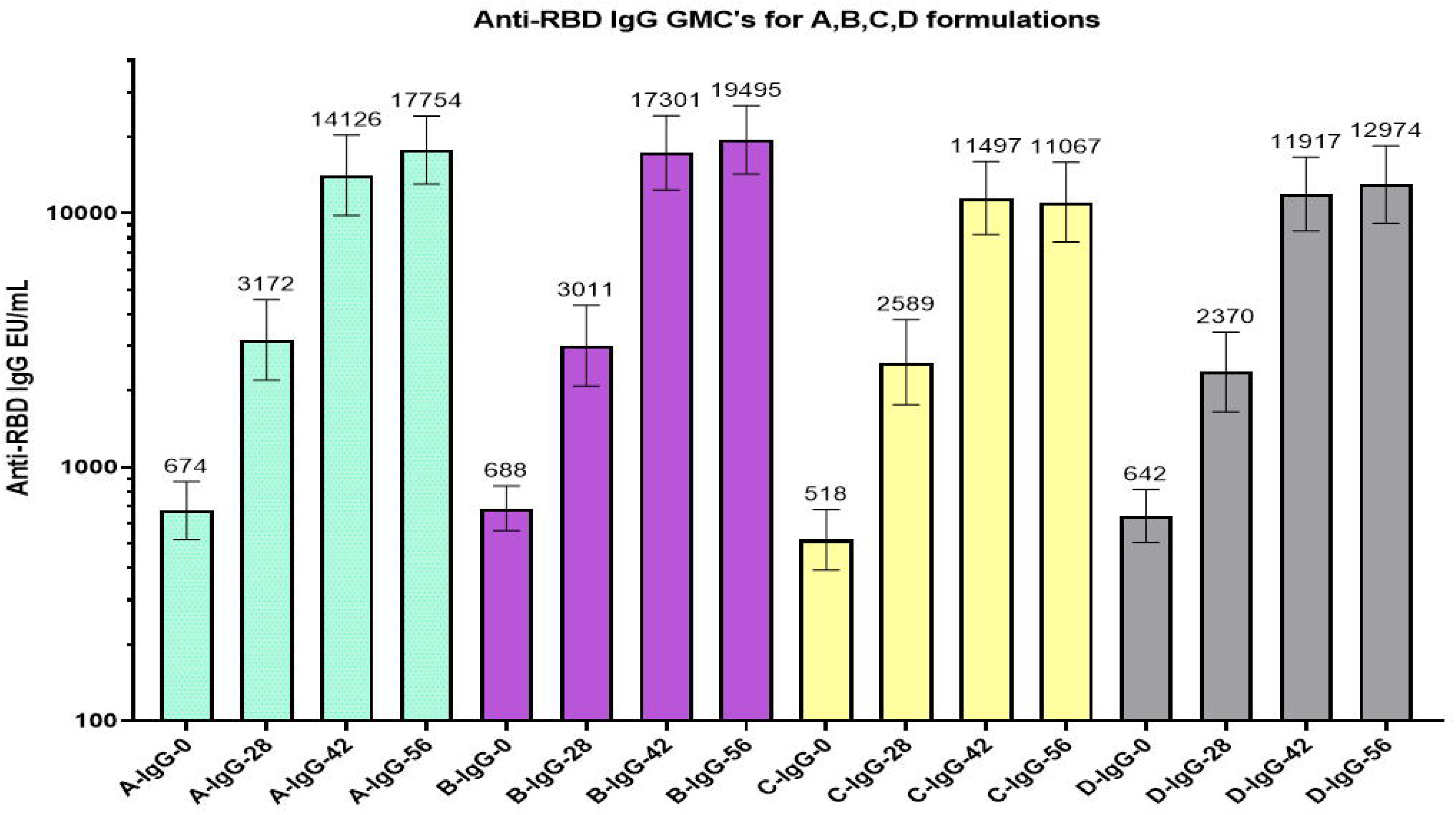
Anti-RBD IgG GMC’s for all four formulation cohorts at Day-0 (prevaccination), Day-28 (Post first-dose), Day-42 & 56 (post second-dose) time-points. GMC”s (top of each column) with 95% Confidence Interval (two-sided bars) are included in the figure.

**Figure 4:**
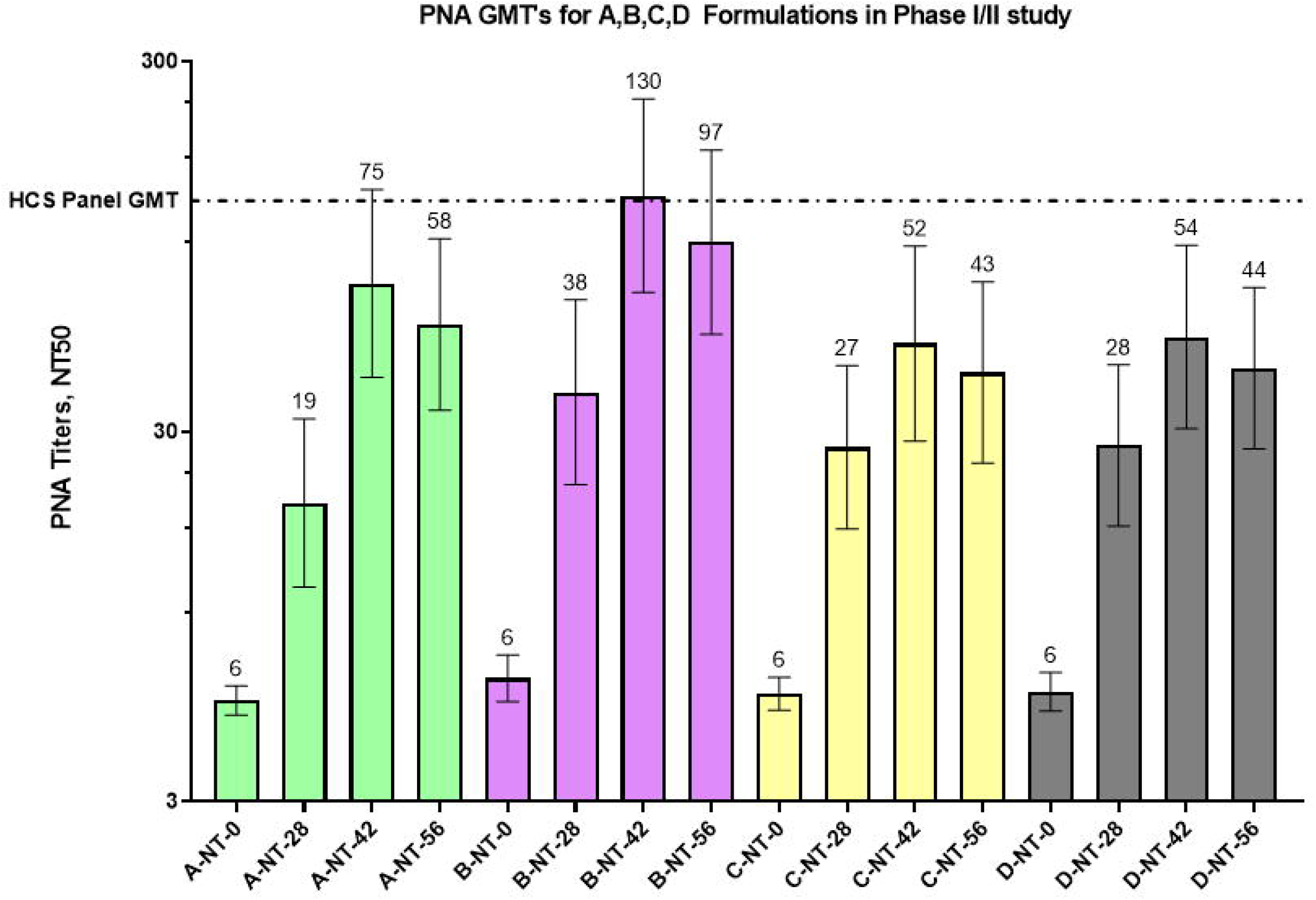
nAb titers by PNA method for all four formulation cohorts at Day-0 (pre-vaccination), Day-28 (Post first-dose), Day-42 & 56 (post second-dose) tune-points. GMCT’s (top of each column) with 95% Confidence Interval (two-sided bars) are included in the figure. Human Convalescent Sera Panel GMT was determined from 273 subject sera samples. All subjects were RT-PCR positive for SARS-COV-2 infection with varying degrees of disease severity.

**Figure 5:**
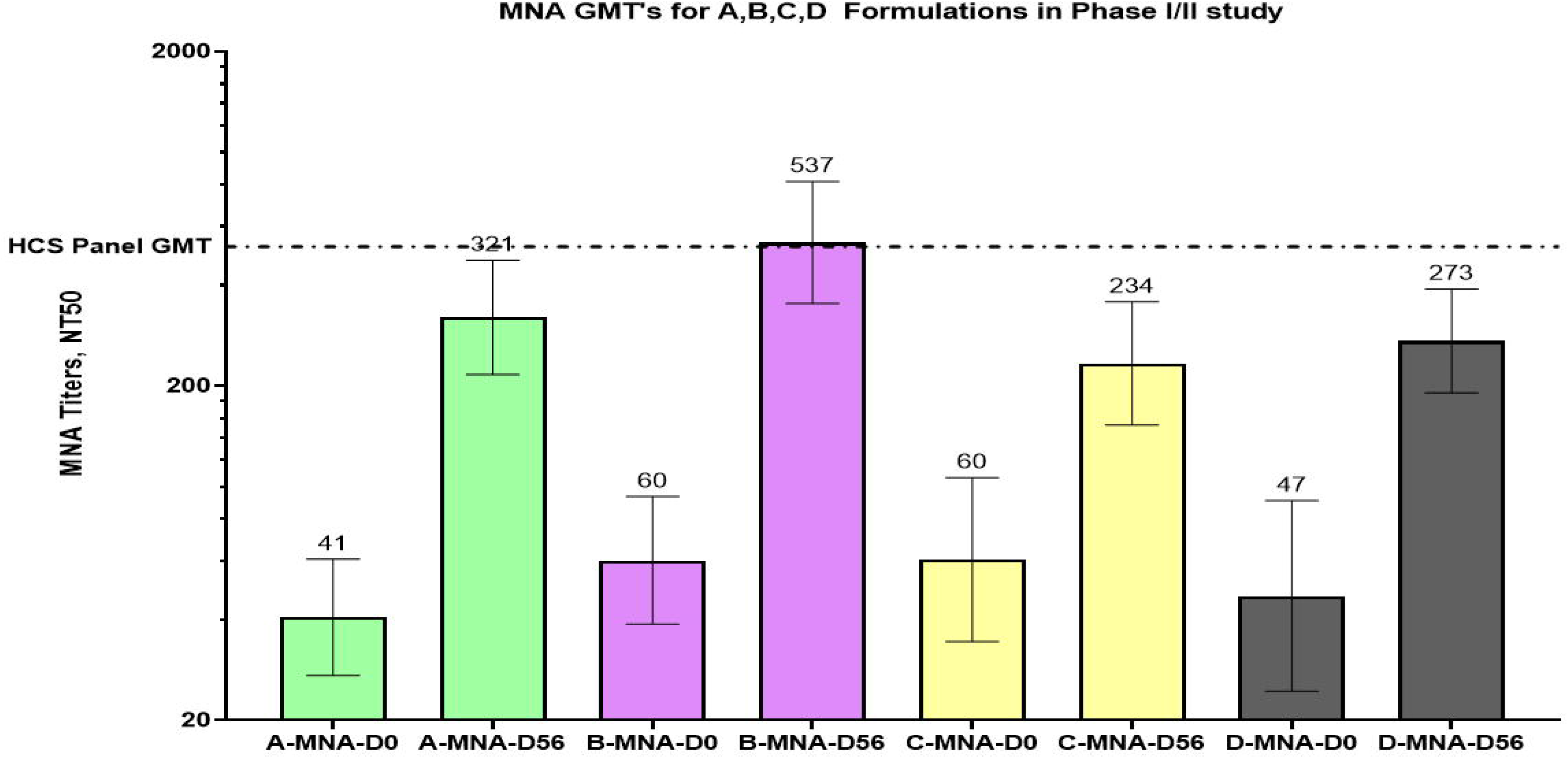
nAb titers by MNA method for all four fo1mulation cohorts at Day-0 (pre-vaccination), Day-56 (post second-dose) time-points. GMT’s (top of each cohunn) with 95% Confidence Interval (two-sided bars) are included in the figure.. Human Convalescent Sera Panel GMT was determined from 32 subject sera samples. All subjects were RT-PCR positive for SARS-COV-2 infection with severe disease. Day-0 titers were measured for only 25% of subjects from each cohort.

**Table 6:** Anti-RBD IgG concentration Geometric Mean Fold Rise from Day0 (pre-vaccination) to Day28 (post first-dose) and to Day42 & 56 (post second-dose) for all four formulation cohorts.

| **Formulation** | **Day28 GMFR** | **Day42 GMFR** | **Day56 GMFR** | **Percentage of Seroconversion** |
| --- | --- | --- | --- | --- |
| **A** | 4.71 | 24.97 | 24.90 | 83% |
| **B** | 4.74 | 25.15 | 28.35 | 90% |
| **C** | 4.58 | 22.03 | 21.21 | 79% |
| **D** | 3.59 | 18.46 | 19.48 | 79% |

GMFR: Geometric mean fold rise, RBD: Receptor binding domain, IgG: Immunoglobulin. Note: Percentage seroconversion observed for Day56 time-point sera samples based on $\geq 4$ -fold rise in anti-RBD-IgG concentration)

**Table 7:** Anti-RBD-IgG1 and IgG4 GMTs for all four formulation cohorts at Day0 (pre-vaccination), and Day56 (post second-dose).

| Formulation | Subjects | IgG1 Titer |  |  | IgG4 Titer |  |  | D56-G1/G4 Ratio<br>GMR |
| --- | --- | --- | --- | --- | --- | --- | --- | --- |
|  |  | D0<br>GMT | D56<br>GMT | GMFR | D0<br>GMT | D56<br>GMT | GMFR |  |
| A | 89 | 74 | 2696 | 36.25 | 31 | 57 | 1.84 | 48.7 |
| B | 90 | 74 | 2940 | 39.7 | 31 | 76 | 2.48 | 37.1 |
| C | 89 | 65 | 1884 | 28.92 | 27 | 50 | 1.81 | 38.0 |
| D | 89 | 89 | 2219 | 24.94 | 32 | 57 | 1.79 | 38.4 |

IgG: Immunoglobulin G, RBD: Receptor binding protein, GMT: Geometric mean titer, D0: Day0, D56: Day56. Note: GMFR for each formulation cohort is calculated from the Geometric Mean for the Fold Rise in titer at Day56 vs Day0 time-point for each cohort. Geometric Mean Ratio was calculated for IgG1 to IgG4 titers at Day56 for all four cohorts.

Neutralizing antibody titers by PNA method where the VSV pseudo virus expressed the spike protein from the Beta strain of SARS-COV-2 were measured in Day56 time-point sera samples from a subset of randomly selected subjects (81 out of 358) for all four cohorts. A comparison of the PNA titers measured against the Ancestral strain (Wuhan, GMT: 362) and the Beta strain (GMT: 161) is shown in Figure-6. For each pair of PNA titers, ratio was calculated and overall Geometric Mean Fold Reduction in PNA titers from Ancestral-Wuhan strain to Beta strain was 2.25.

**Figure 6:**
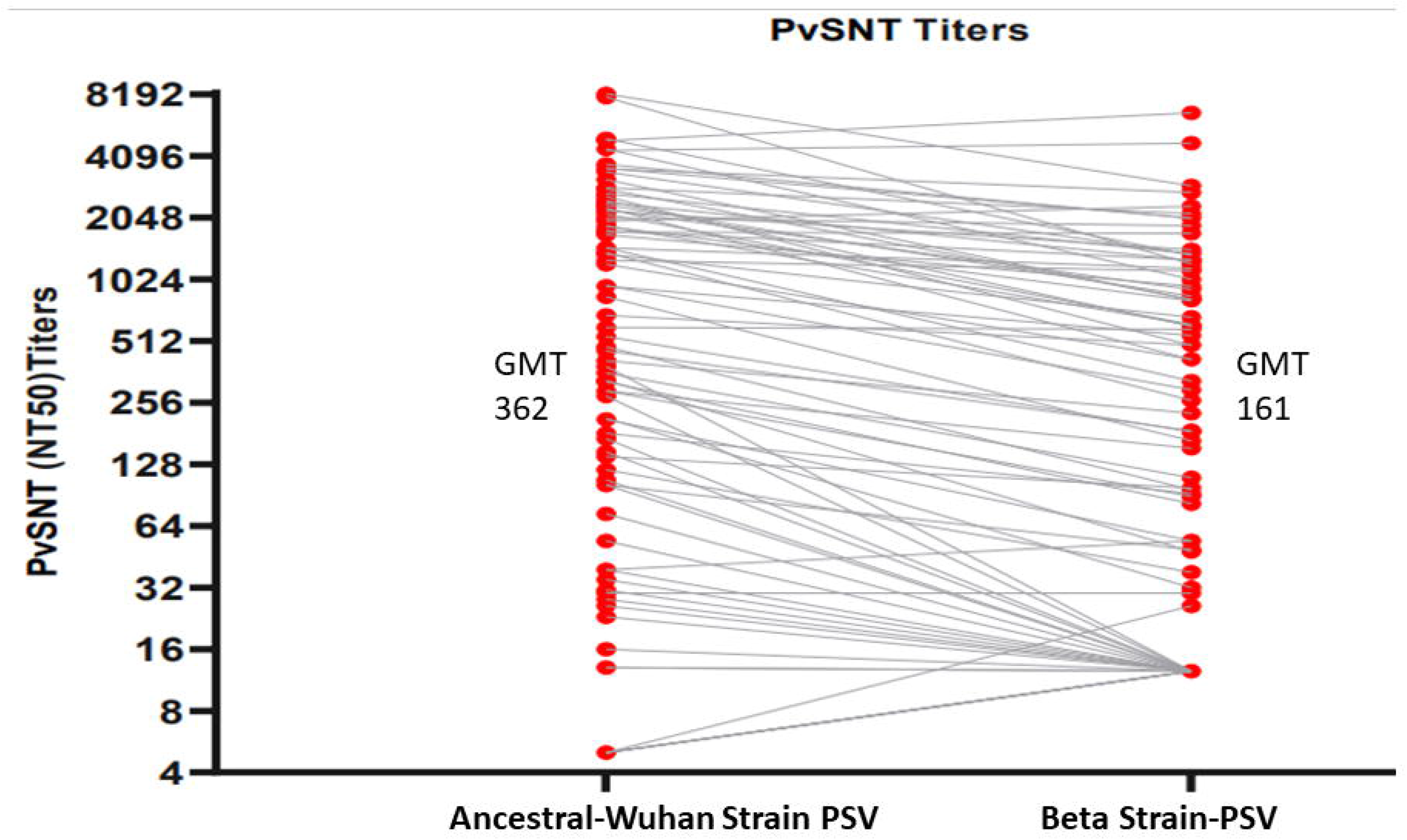
Direct comparison of nAb titers by PNA method against PSV mimicking the Ancestral-Wuhan strain and the Beta strain of SARS-COV-2 for a subset of subjects from all four formulation cohorts at Day-56 time-point (post second-dose). The Geometric Mean Reduction in nAb titers from Ancestral-Wuhan to Beta strain is 2.25 fold.

Cellular immune-response was also assessed from a subset of randomly selected subjects from each cohort. Significantly high IFN-gamma responses were induced by B-formulation (31.4 pg/ml) at Day56 in comparison to others (Table 8).

**Table 8:** Average cytokine concentration at Day0 (pre-vaccination) and Day56 (post second-dose).

| Formulation | Average Interferon-gamma concentration (pg/mL) |  |  |  | Average IL-4 concentration (pg/mL) |  |  |  |
| --- | --- | --- | --- | --- | --- | --- | --- | --- |
|  | D-0 Null | D-0 Active | D-56 Null | D-56 Active | D-0 Null | D-0 Active | D-56 Null | D-56 Active |
| A | 2.24 | 7.95 | 1.75 | 14.01 | 1.45 | 1.63 | 1.31 | 1.36 |
| B | 2.04 | 3.73 | 1.91 | 31.42 | 3.77 | 3.72 | 2.73 | 3.19 |
| C | 1.99 | 5.59 | 2.08 | 22.03 | 1.06 | 1.69 | 1.02 | 1.60 |
| D | 2.59 | 3.02 | 1.87 | 23.03 | 1.07 | 1.23 | 0.89 | 1.22 |

D-0: Day0, D-56: Day56, IL-4: Interleukin 4, pg/mL: picogram per milliliter.
**Note:** Cytokine measured in the supernatants of whole-blood samples incubated in tubes coated with SARS-COV-2 peptides (Active) and without coating (Null) for a subset of subjects from all four cohorts

#### Phase-2 study

The subjects in Phase-2 study received the modified vaccine formulation containing higher CpG1018 adjuvant (Formulation-E, Table 1) to increase robustness and magnitude of the immune-response. Immunogenicity parameters were measured in 100 subjects that were selected based on anti-SARS-COV-2 IgG seronegative status. As the major aim of this immunogenicity analysis was to assess the increase in immune-response due to change in vaccine formulation and hence data is presented for C & B cohorts from Phase-1/2 along with E cohort from Phase-2 trial. Formulation-E induced strong anti-RBD-IgG response (GMC: E-26448 Vs B-17301 and C-11497) compared to C and B, whereas seroconversion rate was comparable in all three formulations (;seroconversion: E-89%; B-88% and C-82%) at day42 (Figure-7 and Table 9). Anti-RBD-IgG1/IgG4 ratio was significantly high in formulation-B (75.4) group compared to formulation-C (38.0) or B (37.1) groups at day42 (Table-10). nAb-titer GMTs measured via PNA method are shown in Figure-8 for Day0 and Day42 time-point with strong NT50 induced by E (GMT: 534) compared to B (GMT: 130) and C (GMT:52). Similarly, formulation-E induced very high nAb-titer GMTs measured via MNA method for Day42 or Day56 time-point (E: 1338 Vs C: 234 Vs B:537) (Figure-9). SARS-COV-2 viruses of Delta and Beta strains were isolated from Indian subjects by THSTI and these virus strains were used in the MNA method of nAb-titer assessment. The MNA GMTs measured against Beta and Delta strains for Day42 sera samples a subset of 20 subjects (Figure-10). Significantly high IFN-gamma responses were induced by E-formulation (99.82pg/ml) at day56 compared to C (22.03pg/ml) and B (31.42pg/ml) (Table 11).

**Figure 7.**
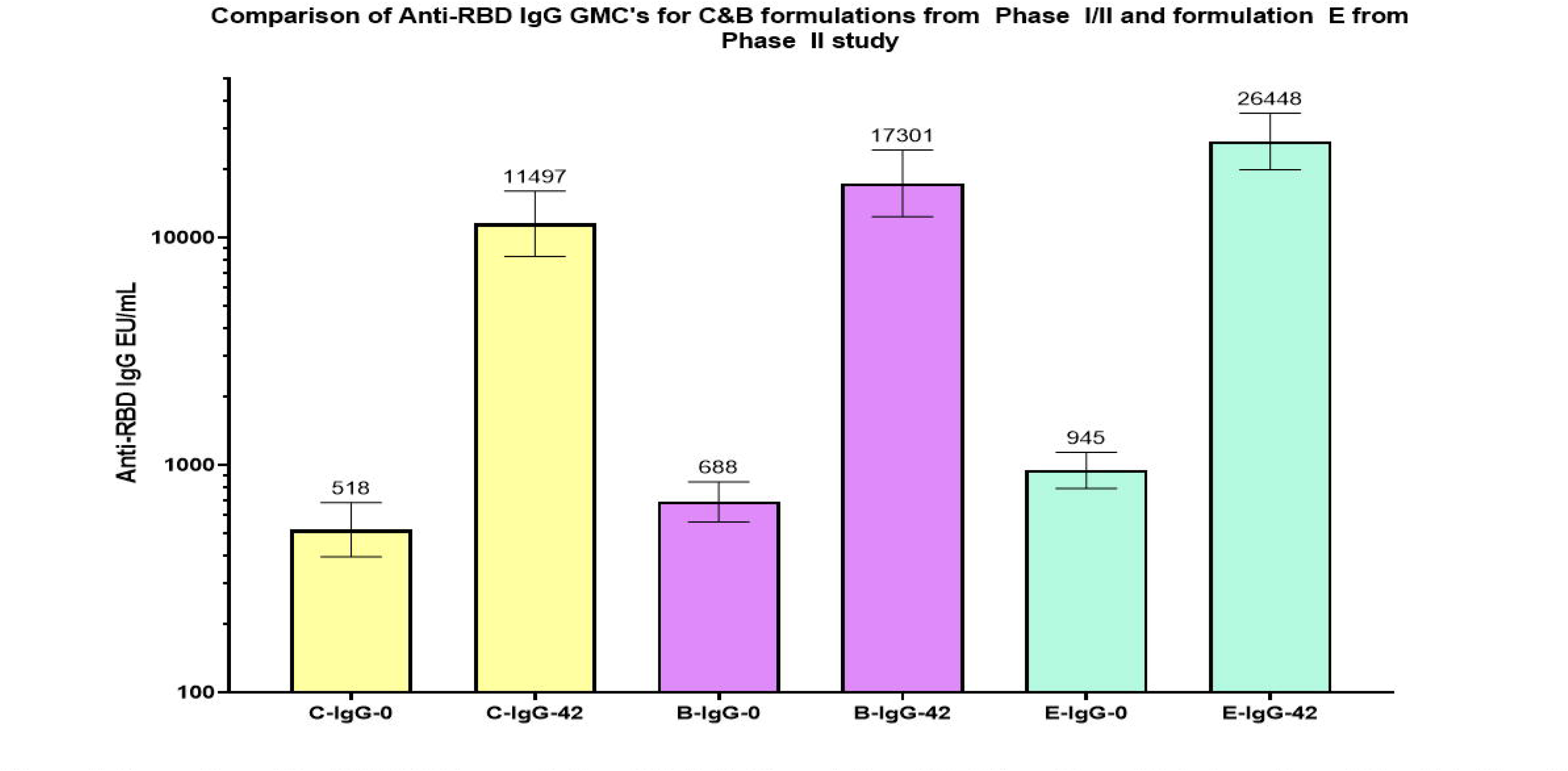
Comparison of Anti-RED IgG concentration GMC’s for Fomulations C & B from Phase I/II study vs. Formulation E in Phase II study. GMC’s are shown at the top of the respective colunms and the 95% Confidence interval is shown as two-sided bars. GMC’s for Day-0 (pre-vaccination) and Day-42 (post two-dose vaccination) are shown in the figure.

**Figure 8.**
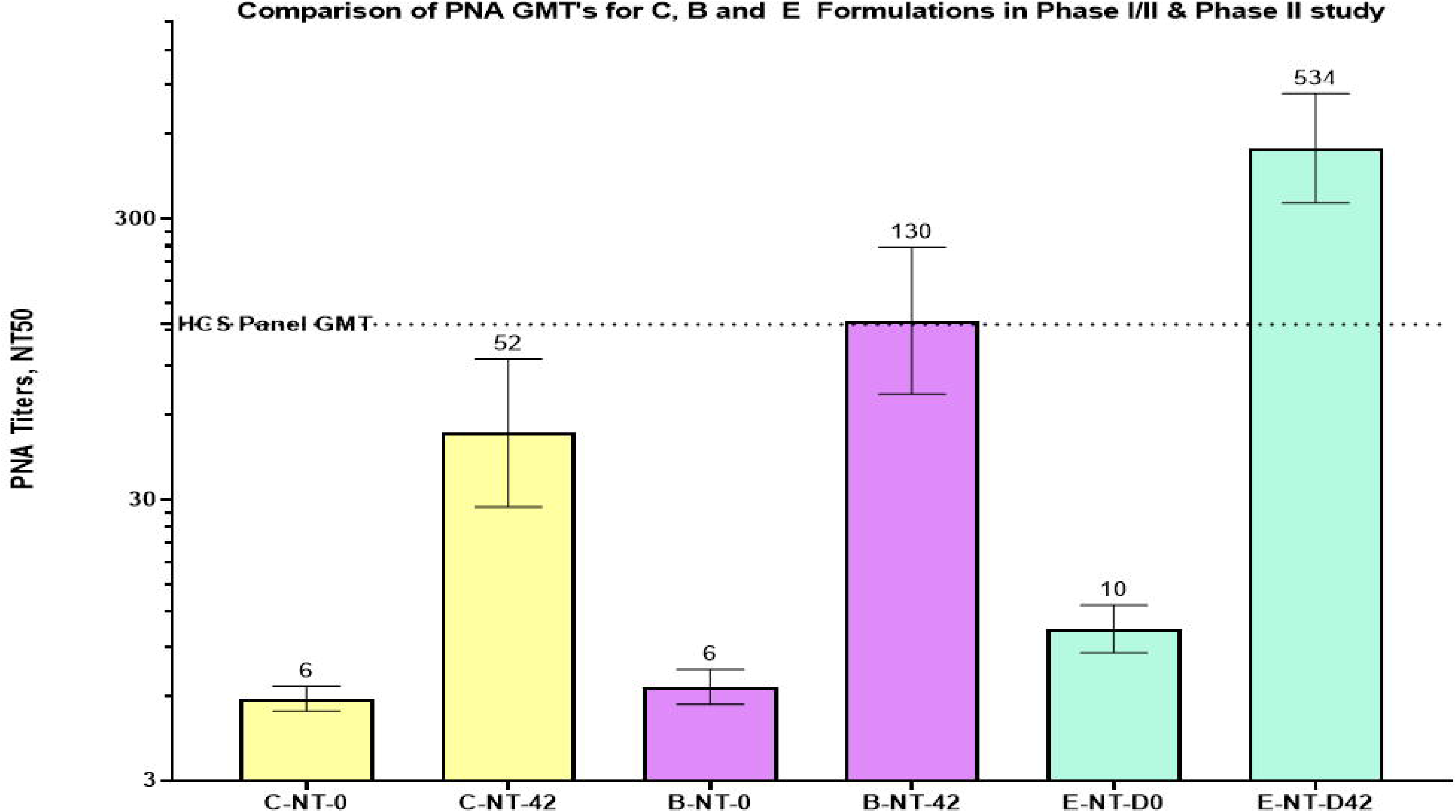
Comparison of nAb titers by PNA method against PSV mimicking the Ancestral-Wuhan strain of SARS-COV-2 for Formulations C & B from Phase I/II study vs. Formulation E in Phase II study. GMT’s are shown at the top of the respective columns and the 95% Confidence interval is shown as two-sided bars. GMT’s for Day-0 (pre-vaccination) and Day-42 (post two-dose vaccination) are shown in the figure.

**Figure 9:**
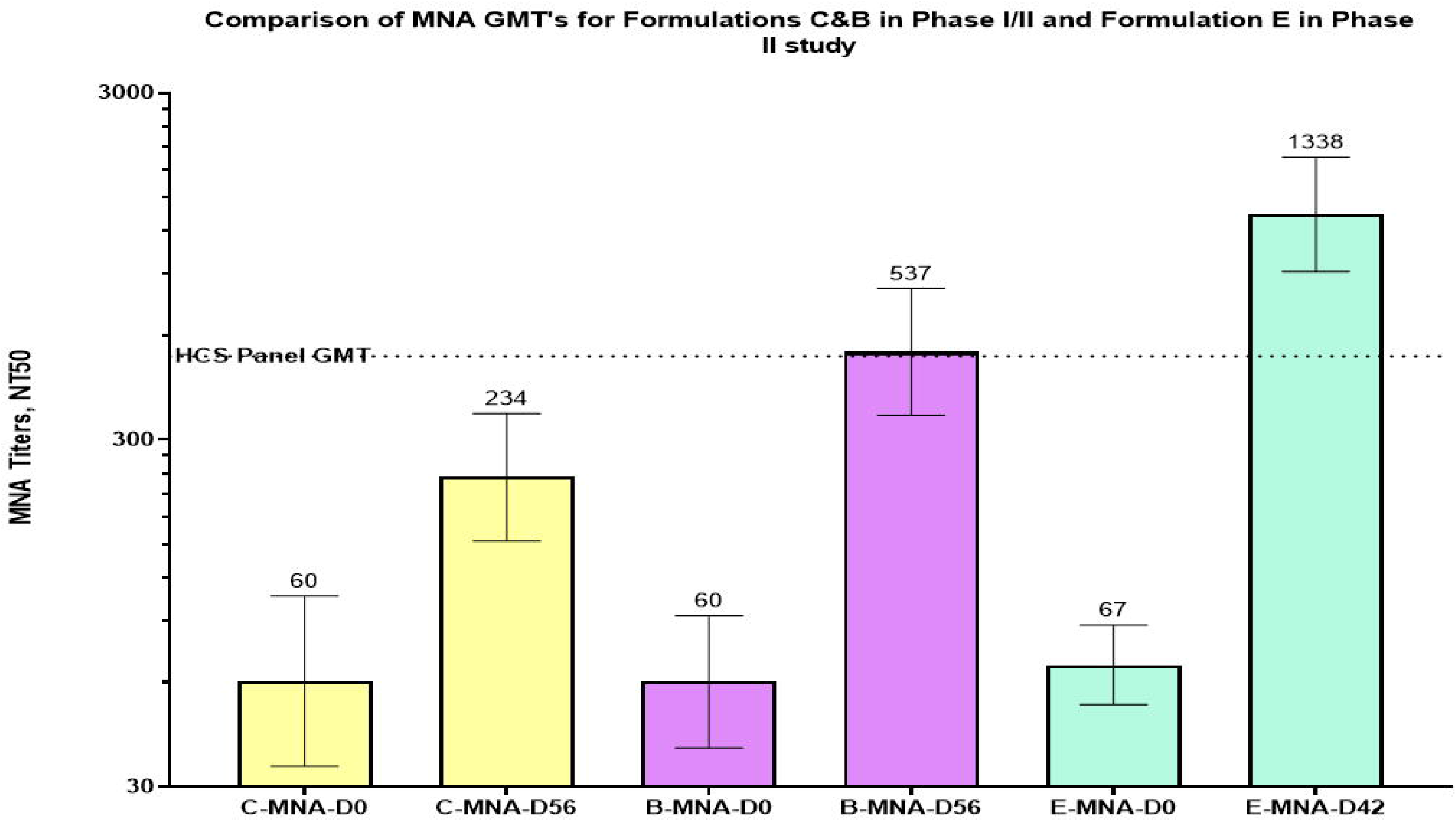
Comparison of nAb titers by MNA method against the Ancestral-Wuhan strain of SARS-COV-2 for Formulations C & B frorn Phase I/II study vs. Formulation E in Phase II study. GMT’s are shown at the top of the respective columns and the 95% Confidence interval is shown as two-sided bars. GMT’s for Day-0 (pre-vaccination) and Day-56 or Day-42 time-point (post two-dose vaccination) are shown in the figure.

**Figure 10:**
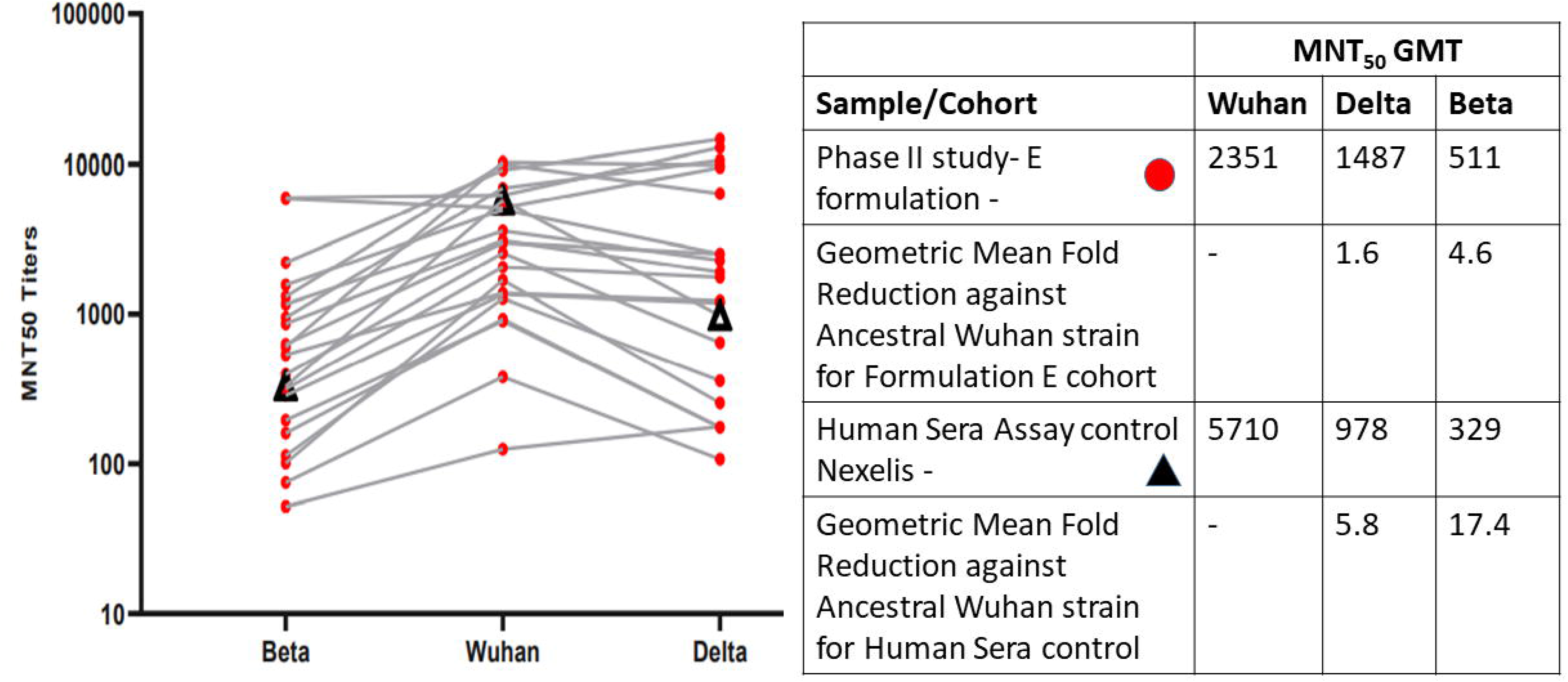
Comparison of nAb titers by PNA method against PSV mimicking the Ancestral-Wuhan strain of SARS-COV-2 for Formulations C & B fron1Phase I/II study vs. Fo1mulation E in Phase II study. GMT’s are shown at the top of the respective columns and the 95% Confidence interval is shown as two-sided bars. GMT’s for Day-0 (pre-vaccination) and Day-42 (post two-dose vaccination) are shown in the figure.

**Table 9:** Anti-RBD IgG concentration Geometric Mean Fold Rise from Day0 (pre-vaccination) to Day42 (post second-dose) for all formulation C&B cohorts from Phase-1/2 and Formulation-E cohort from Phase II study.

| Formulation | Day42 GMFR | % Seroconversion at Day42 |
| --- | --- | --- |
| C | 22.03 | 82% |
| B | 25.15 | 88% |
| E | 27.99 | 89% |

GMFR: Geometric mean fold rise, IgG: Immunoglobulin G. **Note:** Percentage seroconversion observed for Day42 time-point sera samples based on $\geq 4$ -fold rise in anti-RBD-IgG concentration is also reported.

#### Immune-response Persistence data for Phase-1/2 study subjects

The subjects from Phase-1/2 study are planned to be followed for a duration of one year to assess vaccine safety and immune-response. The persistence of immune-response was assessed at 6-months post second-dose i.e.Day208 time-point. All subject sera samples were tested for anti-RBD-IgG concentration and nAb-titers by PNA method at the same laboratories by the same methods. Overall, nAb-titer GMT’s for all the formulations at 6-month time-point were higher than the HCS panel and indicative of high vaccine effectiveness based on comparison with available Correlates of Protection (CoP) data (Figure-11-GMC’s and Figure-12-GMT’s).

**Figure 11:**
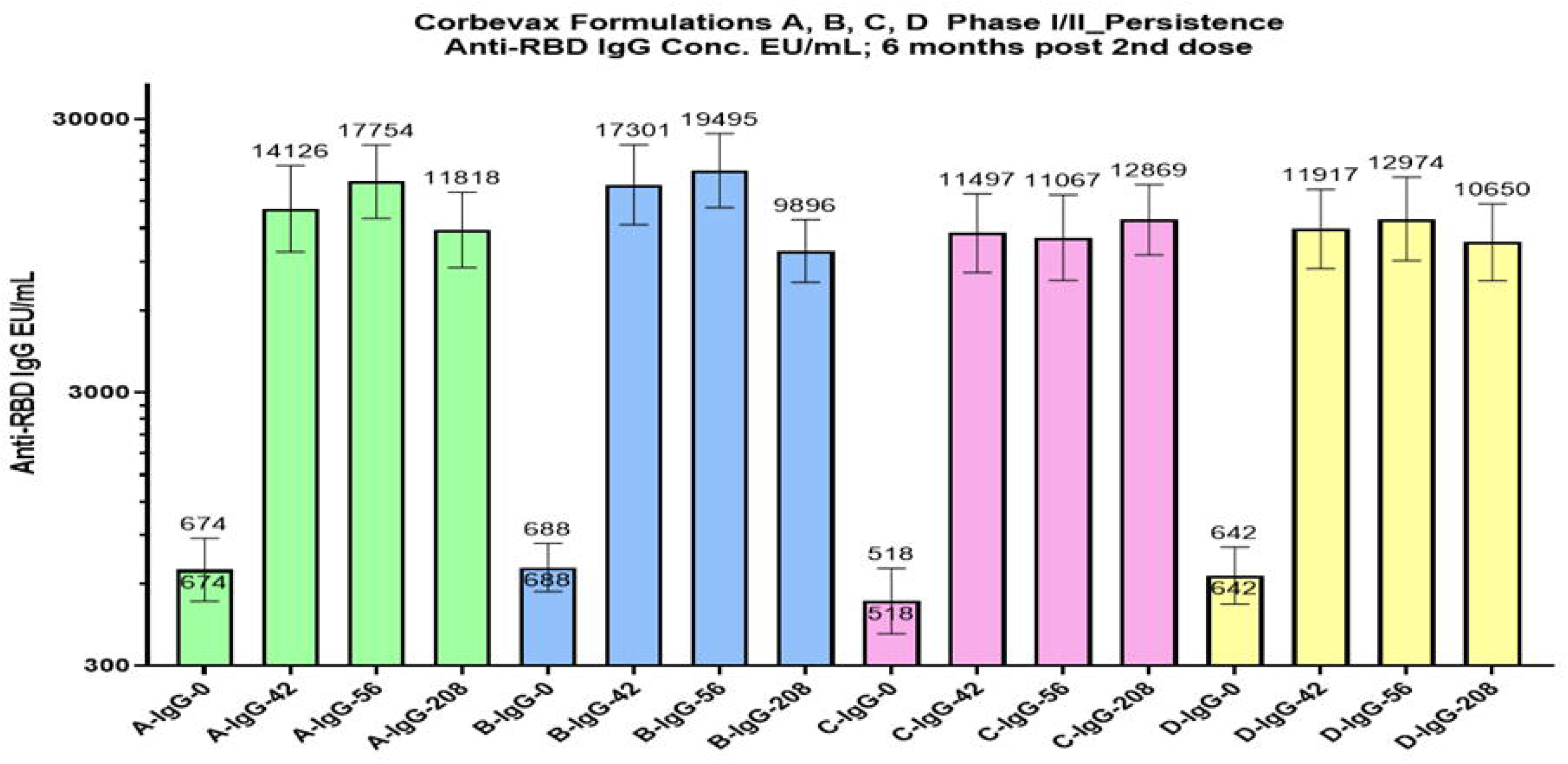
Summary of anti-RBD IgG concentration GMC’s for all four cohorts at Day-0, Day-42, Day-208 time-points

**Figure 12:**
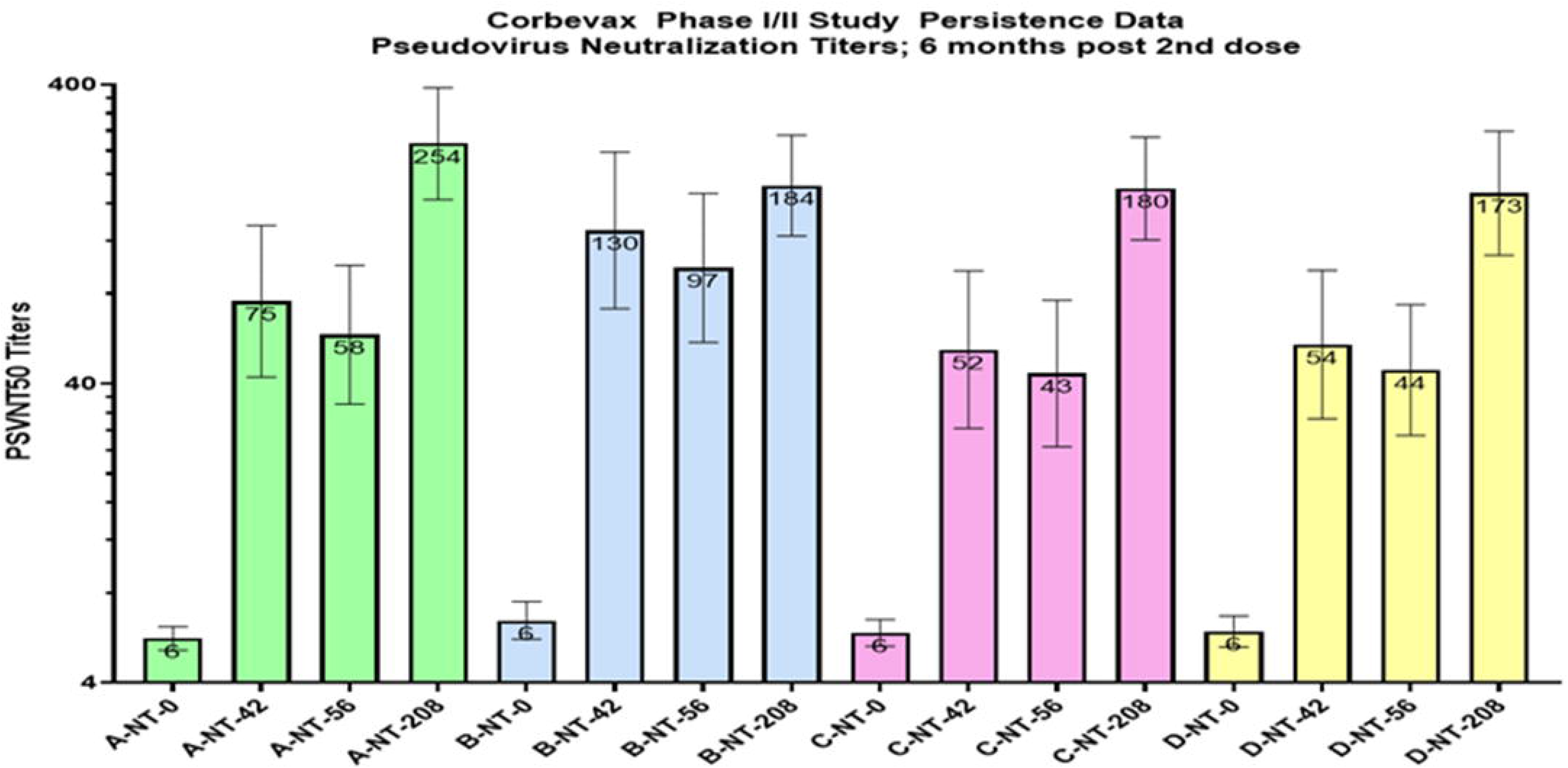
Summary of the GMT’s of nAb titers measured via PNA method for all four cohorts at Day-0, Day-42 and Day-208 time-points

## DISCUSSION

This was a prospective, open-label, Phase-1/2 (randomized) and Phase-2 studies to assess the safety, tolerability, reactogenicity and immunogenicity of four vaccine formulations (Phase-1/2) and optimal formulation (Phase-2) that contained the same antigen i.e. Receptor Binding Domain (RBD) protein, an important target for vaccine development^14^.

### Safety

The vaccine was safe and well tolerated in all formulation cohorts (phase-1/2 and phase-2). Very similar instances of AE’s (the number & percentages of subjects and total number of AE’s) reported after two-dose administrations in phase-1/2 cohorts, whereas slightly higher numbers of AEs reported in the Phase-2 study. Most of the AE’s were mild and no Grade-3 AE’s or Serious AE’s and very low number of MAAE’s were reported in the study. In the ongoing long-term monitoring, other than two cases of mild COVID-19, no additional AE’s were reported in all the cohorts for 10-12 months of phase-1/2 and 5-months of phase-2 study respectively, of monitoring period. Thus, the optimized Corbevax formulation (E) was considered to be safe with minimal reactogenicity to advance into pivotal Phase-3 studies.

### Immunogenicity against Ancestral strain (D614G-Wuhan)

In Phase-1/2 study, both humoral immune-response and cellular immune-response were analyzed at key time-points in the study to determine the impact of various compositions on the overall immune-response.

All four cohorts had similar GMCs at Day0 which increased moderately by Day28 representing low immune-response after first-dose. The GMC’s increased substantially at Day42 and sustained at Day56 showing significant and stable-immune-response post 2nd-dose (Figure-3). Formulation-B had highest immune-response in terms of Anti-RBD GMC’s, %-seroconversion and GMFR’s among all four cohorts.

Antibodies induced by the vaccines are known to neutralize disease-causing agents. All four formulations showed similar trend of increase in nAb-titers i.e., moderate increase after the first dose and significant increase after the second-dose (Day42 & 56 time-points). Similar to the anti-RBD-IgG GMC’s, Formulation-B demonstrate highest GMT’s in the PNA (Day0, 28, 42) and MNA (Day0 and 56) methods and GMFR’s post-vaccination. The post two-dose nAb GMT’s corresponding to Formulation-B i.e., 132 in PNA and 533 in MNA method are higher than established GMT’s of HCS panels in two laboratories viz. GMT of 126 for 273 subjects tested at Nexelis (PNA) and GMT of 522 for 32 subjects with severe COVID-19 disease tested at UKHSA (MNA).

Studies have shown that an appropriate Th1 immune-response can clear the infection and a poor prognosis was associated with cytokine storm that triggers Th2-cells^15, 16^. In this study, anti-RBD-IgG1 (Th1) and IgG4 (Th2) isotype titers were measured. All four formulations showed significant and consistent high ratio of IgG1 to IgG4 titers at Day56 which is a hallmark of skewed Th1-type immune-response. This is thought to be due to the presence of adjuvant CpG1018 in the formulation which is known to skew the immune-response toward Th1 response.

Two key cytokines monitored in the study were IFN-γ (Th1-biased) and Interleukin-4 (Th2–biased). A significant increase was observed in active INF-γ concentration at Day56 compared to baseline samples for all cohorts with highest average concentration observed for the B-formulation. In contrast, only small increase was observed in active IL-4 concentration for Day56 samples for all cohorts. This significant increase in Th1 biased immune-response induced by the Corbevax vaccine further strengthens the role of Th1 responses in fighting against COVID-19 infection^15, 16^.

Significant increase in the magnitude of immune-responses were observed in B-formulations compared to the C-formulation- and the major difference in these two formulations was the adjuvant CpG1018 content i.e., 250 mcg per dose in C-formulation vs. 500 mcg per dose in B-formulation. Hence, to further increase the magnitude and consistency of immune-response, we decided to increase CpG1018 content in the E-formulation to 750 mcg per dose. This increase was compatible with the nature of the formulation i.e., one vial formulation where both RBD protein and adjuvant CpG1018 were almost completely adsorbed to alum, and this became the final optimized formulation of Corbevax vaccine. The Phase-2 study served to confirm the expected improvement in the immune-response parameters due to increase in the CpG1018 adjuvant content while maintaining the excellent safety profile. Significant improvement in anti-RBD-IgG concentrations, anti-RBD-IgG1 titers and nAb-titers as measured by PNA and MNA method was observed for E-formulation vs the B-formulation which confirmed the enhancement of immune-response via inclusion of higher CpG1018 content in the vaccine formulation. Cellular immune-response was also higher for E-formulation vs the B-formulation. These comparisons are summarized in Figures 7, 8 and 9 and Tables 10 and 11.

**Table 10:**
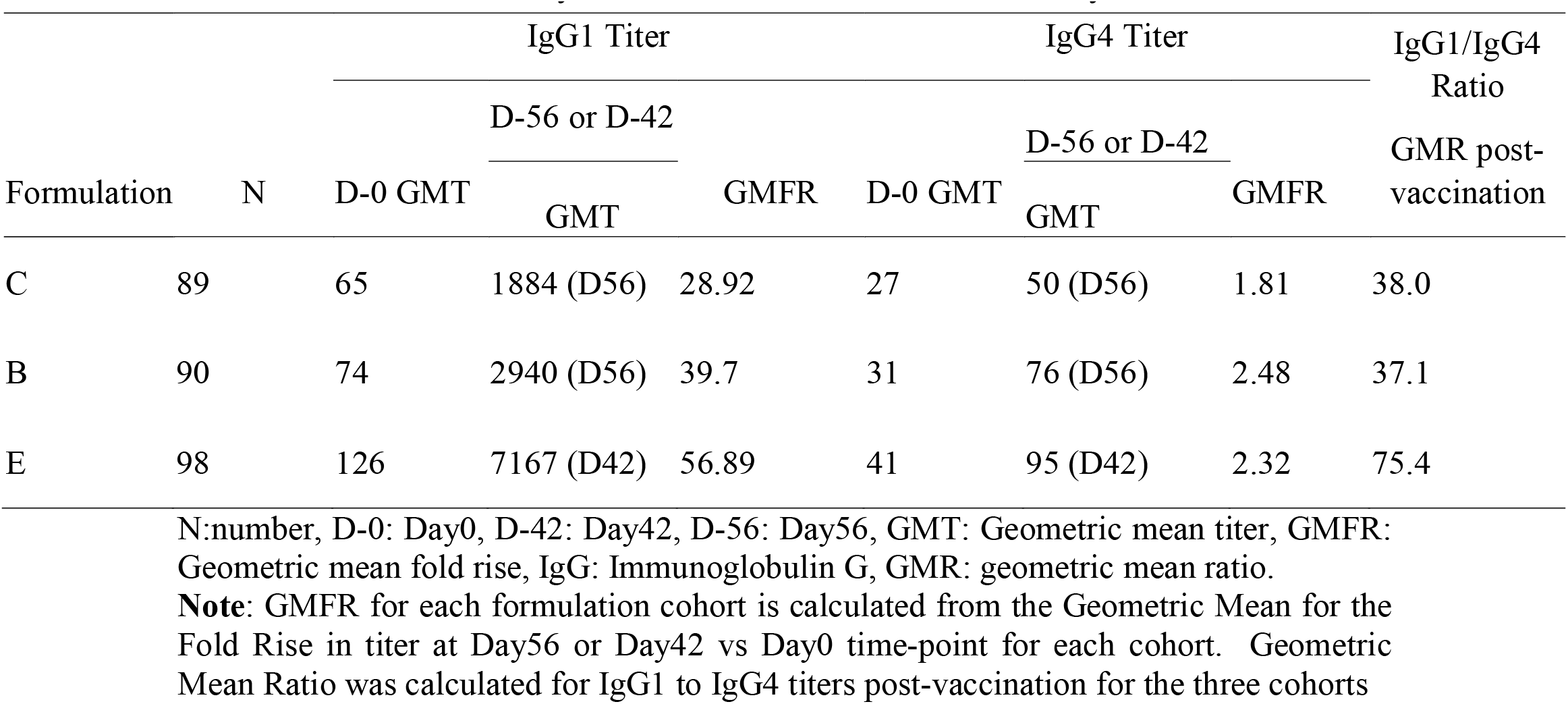
Comparison of Anti-RBD-IgG1 and IgG4. GMTs for all Formulation C&B cohorts from Phase-1/2 study and Formulation E from Phase-2 study.

**Table 11:** Average cytokine concentration at Day0 (pre-vaccination) and Day56 or Day42 (post second-dose).

| Formulation | Average Interferon-gamma concentration<br>(pg/mL) |  |  |  | Average IL-4 concentration<br>(pg/mL) |  |  |  |
| --- | --- | --- | --- | --- | --- | --- | --- | --- |
|  | D-0 Null | D-0 Active | D-56 Null | D-56 Active | D-0 Null | D-0 Active | D-56 Null | D-56 Active |
| C | 1.99 | 5.59 | 2.08 | 22.03 | 1.06 | 1.69 | 1.02 | 1.60 |
| B | 2.04 | 3.73 | 1.91 | 31.42 | 3.77 | 3.72 | 2.73 | 3.19 |
| E | 7.03 | 26.24 | 2.63 | 99.82 | 5.19 | 4.26 | 8.10 | 10.96 |

D-0: Day0, D-56: Day56, IL-4: Interleukin 4, pg/mL: picograms per milliliter. **Note:** The cytokine concentrations are measured in the supernatants of whole-blood samples incubated in tubes coated with SARS-COV-2 peptides (Active) and without coating (Null) for a subset of subjects from Formulation C&B cohorts from Phase-1/2 study and Formulation-E cohort from Phase-2 study.

### Cross-neutralization of Variants of Concern

One of the most important performance attributes of COVID-19 vaccine is its ability to provide protection against infection by variant strains as measured by nAb-titers against the key variants. At the time of Phase-1/2 study execution, the most important Variant of Concern was the Beta strain. Nexelis laboratory created a Pseudovirus that expressed the spike protein of the Beta strain of SARS-COV-2 with all the known mutations and this same PSV strain was used in the PNA method to assess the nAb-titers against Beta-PSV. In a subset of 81 subjects across all four cohorts, nAb-titers against the Beta-PSV were measured and compared with the observed nAb-titers against the Ancestral D614G-PSV strain. This comparison (Figure-10) shows that the overall fold reduction of only 2.25-fold in nAb-titers against the Beta-strain in comparison to Ancestral strain. This cross-neutralization potential of Corbevax formulation is significantly superior to other vaccines (e.g., mRNA, adenovector vaccines) which showed 5-10-fold reduction in nAb-titers from Ancestral to Beta-strain^17, 18^

In Phase-2 study, a subset of subject sera samples was tested against wild-type Beta and Delta-strains of SARS-COV-2 isolated in India at THSTI laboratory. This comparison (Figure-10) showed only 1.6-fold reduction in nAb GMT’s from Ancestral strain to Delta strain and 4.6-fold reduction in nAb GMT’s from Ancestral strain to Beta strain. More importantly, all twenty subject sera samples demonstrated detectable-nAb-titers against both Beta and Delta-strains. The corresponding convalescent sera control (obtained during the initial wave i.e., infection from the ancestral strain) showed 5.8- and 17.4-fold reduction in nAb-titers against Delta and Beta-strains respectively in the same assay. Thus, Corbevax vaccination yields the most consistent cross-protection against two most relevant VOC’s and this protection potential is significantly superior to other vaccines.

### Persistence of Immune-response

Longevity of immune-response is very important attribute of any vaccine and that is routinely assessed during long-term monitoring. The subjects in Phase-1/2 study are part of long-term monitoring and immunogenicity, data is available till 6^th^ month post second-dose. Minimal change in anti-RBD-IgG GMC’s and nAb GMT’s at 6 months post second-dose of the vaccine as compared to 2&4-week time-point after the second-dose (Figure-10 and 11). This shows excellent persistence of humoral immune-response over a significant duration of 6-months post-vaccination. This attribute of Corbevax is significantly superior to other vaccines that have demonstrated 70-90% drop in binding-antibody and nAb-titers in the same duration^19–21^. During the approximately 1-year of monitoring period post two-dose vaccination from Phase-1/2 study, only two subjects (one each from C and D formulation cohorts) reported mild symptomatic COVID-19 infection which corresponds to COVID-19 incidence rate of approximately seven cases per 1000 person-years which corresponds to very high vaccine effectiveness.

### Expected Corbevax effectiveness in preventing symptomatic infection from Ancestral strain

Based on the analysis conducted during Phase-3, efficacy studies was part of product approval for Spikevax^22^ (Moderna Inc;) and Vaxzveria^23^ (AstraZeneca Inc;); nAb-titers in sera samples after two-dose vaccine administration were observed to correlate with the protection against symptomatic COVID-19 infection. Both studies also reported this CoP information in terms of nAb-titers expressed in IU/mL by using calibration factors to convert the study specific nAb-assay titers to the WHO-International Standard. This CoP evaluation showed that if nAb GMT’s of >100 IU/mL post two-dose vaccination correspond to significant vaccine efficacy viz. >90% higher than placebo-control. The nAb GMT’s at Day-42 time-point (14-days after second-dose, same as that used for Spikevax and Vaxzveria) in Corbevax Phase-2 study were 285 IU/mL based on PNA method and 329 IU/mL based on MNA method which is indicative of vaccine effectiveness of >90%. There are reports which showed that the ratio of nAb GMT’s post-vaccination to the HCS panel nAb GMT also correlates with vaccine efficacy independent of the nature of COVID-19 vaccine^24^. These ratios post Corbevax vaccination in Phase-2 trial were calculated as 4.2 and 2.6 based on PNA and MNA method respectively and these ratios also indicate >90% vaccine efficacy.

## CONCLUSIONS

From the present study it is clear that Biological E’s Corbevax vaccine, was safe and well tolerated in healthy adult volunteers of Indian origin aged 18-55 years with no AE’s of clinical concern. Comparison of the nAb GMTs observed post two-dose Corbevax vaccination in Phase-2 study with the CoP evaluations and vaccine efficacy data for other COVID-19 vaccines indicate, vaccine efficacy of >90% for protection against symptomatic COVID-19 infection. Excellent maintenance of binding antibody and nAb-titers over 6-months duration post two-dose vaccination from all four vaccine formulations also indicate that the high level of protection from symptomatic infection will be sustained for extended duration as opposed to the significant waning of immune-response and vaccine effectiveness observed for most of the other COVID-19 vaccines. Based on the excellent safety profile, significant and robust humoral and cellular immune-response and desired Th1 skewed immune-response post Corbevax vaccination, pivotal Phase-3 clinical trials have been initiated.

### Study limitations

The present study is an open-label study and not a randomized double-blind study. The study population did not include pediatric and elderly age group (65+Years). Immune-response data for the optimized formulation is available till two weeks post second-dose and longevity of the immune-response for the optimum formulation will be available in the future during long-term monitoring. The subject cohorts were limited in number in Phase-1/2 and Phase-2 studies and the vaccine performance in larger cohorts in wider age-groups (5-80) will be assessed in the ongoing Phase-3 studies.

## Data Availability

Additional study data which is not part of the manuscript can be made available upon request and addressed to the corresponding author Dr. Subhash Thuluva at his.

## Acknowledgments

We are thankful to all the study participants, the principal investigators, and the study staff at all the clinical sites. In addition, we are thankful to the team at Dang’s Lab, New Delhi, led by Dr. Leena Chatterjee, Dr Arjun Dang, Mr Dinesh Kuma and Mr. Shakeeb Mohammad, performed the functions of study sample coordination (receipt, accessioning, aliquoting, labeling, storage, and dispatch) as well as conducted testing of all the ELISA assays (anti-RBD and cytokines) for all the samples. We are also thankful to the team (Mr Anantharaj, Mr Kamal Pargai, Mr Parveen Kumar, Mr Alok Tripathi, Ms Neha Garg, and Mr Shamsher Dhull) at THSTI, led by Dr. Guruprasad Medigeshi, who conducted neutralizing antibody titer testing of all the sera samples against SARS-COV-2 strains. Finally, we appreciate the help rendered by the team at Nexelis, led by Dr. Luc Gagnon and manager Mary Osei-Twum in conducting the neutralizing antibody titer testing of all the sera samples against pseudo virus expressing SARS-COV-2 spike protein.

Dr. Maria Bottazzi and Dr. Peter Hotez, and their scientific team at the Center for Vaccine Development at Baylor College of Medicine/Texas Children’s Hospital, created and produced the recombinant Pichia Pastoris strain expressing the RBD protein. Dynavax, Inc supplied the adjuvant CpG1018 used in the Corbevax formulations. The clinical assay development team led by Dr. Arun Kumar at CEPI helped with neutralizing antibody titer assays in the supply of reagents and establishing assay consistency across multiple laboratories. The writing support for the manuscript was provided by Syneos® Health. Authors would like to thank Dr. Suneetha Pothakamuri and Mr. Kamal Thammireddy for their valuable support in reviewing and finalizing the manuscript. Development of this vaccine candidate would not have been possible without the efforts of manufacturing, quality control, quality assurance and regulatory teams from Biological E. The authors would like to thank Scientific Advisory Board (SAB) and management of Biological E Limited for their support and valuable guidance. All authors wish to express their appreciation and gratitude for all front-line healthcare workers.

