## Supplementary Information for "Selection of optimum formulation of RBD-based protein sub-unit covid19 vaccine (Corbevax) based on safety and immunogenicity in an open-label, randomized Phase-1 and 2 clinical studies"

**Supplementary Material**

**Inclusion Criteria**

Subjects were enrolled in the study based on the following inclusion criteria.

1. Participants of either gender between ≥18 to ≤55 years of age at phase-I and ≥18 to ≤65 years of age at phase-II at the time of 1^st^ vaccination.
2. Participants virologically seronegative to SARS-CoV-2 infection by RT-PCR and anti-SARS-CoV-2 antibody prior to enrolment.
3. Participants seronegative to HIV 1 & 2, HBV and HCV infection prior to enrolment.
4. Participants considered of stable health as judged by the investigator, determined by medical history and physical examination with normal vital signs as defined in the protocol. *[Normal vital signs defined as pulse rate of ≥60 to ≤100 bpm; blood pressure systolic of ≥90 mm Hg and <140 mm Hg; diastolic ≥ 60 mm Hg and <90 mm Hg; body temperature <100.4ºF prior to enrolment].*
5. Female participants of childbearing potential negative to urine pregnancy test and willingness to avoid becoming pregnant through use of an effective method of contraception or abstinence from the time of study enrolment until six weeks after the last dose of vaccination.
6. Agrees not to participate in another clinical trial at any time during the total study period.
7. Agrees to refrain from blood donation during the course of the study.
8. Agrees to remain in the town where the study centre is located, for the entire duration of the study.
9. Willing to allow storage and future use of collected biological samples for future research in an anonymised form.

**Exclusion Criteria**

Subjects were excluded from the study based on the following exclusion criteria:

1. History of vaccination with any investigational vaccine against COVID-19 disease.
2. Seropositive to IgG antibodies against SARS CoV-2
3. Living in the same household of any COVID-19 positive person.
4. Pregnant women, nursing women or women of childbearing potential who are not actively avoiding pregnancy during clinical trials.
5. Seriously overweight (BMI ≥ 40 Kg/m2).
6. Use of any investigational or non-registered product other than the study vaccine during the trial period or 3 months prior to enrolment.
7. History of receipt of any licensed vaccine within 1 month prior to screening, likely to impact on interpretation of the trial data (e.g., influenza vaccines);
8. Current or planned participation in prophylactic drug trials for the duration of the study.
9. Any clinically significant abnormal haematology and biochemical laboratory parameters tested at screening as judged by the investigator.
10. Body temperature of ≥100.4°F (>38.0°C) or symptoms of an acute illness at the time of screening or prior to vaccination.
11. History of severe psychiatric conditions likely to affect participation in the study.
12. History of any bleeding disorder (e.g., factor deficiency, coagulopathy, or platelet disorder).
13. History of allergic disease or reactions likely to be exacerbated by any component of the Biological E’s four COVID-19 vaccine formulations.
14. Chronic respiratory diseases, including asthma.
15. Chronic cardiovascular disease, gastrointestinal disease, liver disease, renal disease, endocrine disorder, and neurological illness.
16. Any other serious chronic illness requiring hospital specialist supervision.
17. Suspected or known current alcohol abuse as defined by an alcohol intake of greater than 42 units every week for at least one year.
18. Chronic administration (defined as more than 14 days in total) of immunosuppressant (e.g., corticosteroids, cytotoxic drugs or antimetabolites, etc.) or other immune-modifying drugs (e.g., interferons) during the period starting six months prior to the first vaccine dose including use of any blood products. For corticosteroids, this will mean prednisone ≥0.5 mg/kg/day, or equivalent. Inhaled and topical steroids are allowed.
19. Any confirmed or suspected immunosuppressive or immunodeficient condition, based on medical history and physical examination (no laboratory testing required).
20. Any medical condition that in the judgment of the investigator would make study participation unsafe.
21. Individuals who are part of the study team or close family members of individuals conducting the study.

**Methodology**

As per the kit manufacturer, subjects with antibody concentrations below 12 Antibody Units/mL were designated as sero-negative and were selected in the trial (Diasorin kit). Health status assessed during the screening period was based on medical history and clinical laboratory findings, vital signs, and physical examination. All those who were part of any other clinical trial, with a history of vaccination with any investigational vaccine against Covid-19 disease, BMI ≥ 40 Kg/m^2^, any other health issues, or were on immunosuppressants, immunodeficient conditions or sero-positive for SARS-COV-2 were excluded from the study.

**Procedure**

The number and percentage of subjects with Adverse events (AEs) and severe adverse events (SAEs) were presented overall by system organ class (SOC) & preferred term (PT). The percentage of subjects with at least one local AE (solicited and unsolicited), with at least one systemic AE (solicited and unsolicited) and with any AE during the solicited follow-up period were tabulated with exact 95% confidence interval (CI). The same calculations were performed for symptoms rated as Grade 3 and above. Systemic and local tolerability, recorded in subject diaries, were summarized in a frequency table with percentages based on the number of observed values. Serious adverse events, related AEs, AEs leading to death or withdrawal, solicited AEs, and MAAEs were summarized separately.

**Immunogenicity Analysis**

Humoral immune responses were evaluated by following methods:

1. Anti-RBD Antibody response: Anti-RBD IgG concentration in the subject sera samples were measured at pre-vaccination (Day-0), post first dose (Day-28) and at multiple time-points post second dose (Day-42, Day-56, Day-208) using a validated enzyme linked immunosorbent assay (ELISA) method executed at Dang’s Lab, New Delhi, India. A monoclonal antibody, CR-3022 supplied by Lake Pharma Inc, CA, USA, that binds specifically to the RBD protein was used to generate standard curve of ELISA OD response vs. antibody concentrations. ELISA concentration equivalent to 1 ng/mL of CR-3022 antibody binding concentration was assigned concentration of 1 anti-RBD ELISA Unit/mL and thus anti-RBD IgG concentration in the sera samples were reported in EU/mL. The National Institute for Biological Standards and Control, UK plasma reference standard 20/130 was used as a positive control on all the plates with a control range of (**8151** to **15137** EU/mL) for validity consideration^1^. Geometric means were calculated for various subject cohorts at specific time-points and fold rise in anti-RBD concentrations for all time-points post start of vaccination were calculated in relation to the pre-vaccination concentrations and then geometric mean fold rise (GMFR) were calculated for each cohort.
2. Anti-RBD IgG subclass response: Anti-RBD IgG1 and IgG4 subclass titers were measured in the sera samples at pre-vaccination (Day-0), post first dose (Day-28) and at multiple time-points post second dose (Day-42, Day-56) using a validated ELISA method executed at Dang’s Lab, New Delhi, India. A pool of sera was prepared with very low anti-RBD IgG concentration from pre-vaccination samples and the threshold ELISA OD for assignment of titer was calculated as threshold OD = Average of blank pool OD + 5.56*standard deviation. Each sera sample was tested as a series of two-fold dilutions (starting from 50-Fold) and the dilution that resulted in OD higher than the threshold OD was assigned as the IgG1 or IgG4 titer. Geometric means were calculated for various subject cohorts at specific time-points and fold rise and GMFR in anti-RBD IgG1 and IgG4 titers for all time-points post vaccination were calculated in relation to the pre-vaccination titers.
3. SARS-COV-2 Virus Neutralization: SARS-COV-2 neutralizing antibody titers (nAb titers) were measured via 2 different methods^2^. Microneutralization Assay (MNA) was employed using Wild-Type SARS-COV-2 strain (Victoria isolate 01/2020). Pseudo virus Neutralization Assay was employed using Vascular Stomatis Virus expressing the SARS-COV-2 Spike protein and reporter gene Luciferase. The MNA was conducted at Translational Health Science and Technology Institute (THSTI), Faridabad, India and PNA was conducted at NEXELIS, Quebec, Canada; both are Coalition for Epidemic Preparedness Innovation (CEPI-network) labs. The nAb testing was conducted as per methods described previously^2^. Conversion factors have been established by both the laboratories to enable conversion of the Neutralization Titer (NT_50_) values to WHO-International Standard (NIBSC-20/136) and report the (NT_50_ values in International Units/mL^3^. MNA values were divided by 4.064 and PNA values were divided by 1.875 to obtain the titers in IU/mL when required for comparison. Geometric mean titers were calculated at scheduled time-points and fold Rise from the pre-vaccination values were calculated along with GMFR. Sera samples that did not demonstrate minimum 50% neutralization of the virus at the initial dilution i.e. the of the assay, titers were assigned as LLOQ/2. For key GMT/GMC values, 95% Confidence Intervals (95%CI) were also calculated. All the subject sera samples were tested for nAb titers using PNA method in Phase I/II and Phase II study. The subjects that had PNA titers below the LLOQ were considered as sero-negative for analysis while subjects with PNA titers above LLOQ were considered as sero-positive for analysis. Seroconversion rate for anti-RBD IgG concentration and nAb titers were calculated as follows: For sero-negative subjects, ≥4‑fold rise in IgG or nAb titers was considered to be seroconverted while for sero-positive subjects, ≥2‑fold rise in IgG or nAb titers was considered to be seroconverted.
4. Cellular immune responses were assessed in a subset of subjects in terms of cytokine secretion in TrueCulture^®^ tubes^4^. Whole blood samples from the subjects were incubated in growth medium in TrueCulture^®^ tubes coated with SARS-COV-2 Peptides (supplied by Myriad Inc, Texas, USA). The Peripheral Blood Mononuclear Cells present in the blood are stimulated by the peptides that resemble the vaccine antigen and in response to the stimulation the cells produce various cytokines. The culture supernatants were recovered and 2 cytokines; Interferon-gamma and Interlukin-4, secreted in the supernatant were measured by standard ELISA kits supplied by Becton Dickinson, USA. The whole blood stimulation was done at the clinical trial sites and the cytokine ELISA was conducted at Dang’s Lab. Cytokine values in pictogram/mL were reported for the sera samples and arithmetic averages were calculated for each cohort.
5. **Institutional Ethics Committees (IECs) of 5 study sites for phase-1/2 (BECT062) study**

| **Name of Committee** | **Approval Status** | **Date of Approval** |
| --- | --- | --- |
| Ethics Committee, St. Theresa’s Hospital, Hyderabad | Approved | 10/11/2020 |
| Guru Teg Bahadur Hospital Ethics Committee, Delhi | Approved | 28/11/2020 |
| IEC, All India Institute of Medical Sciences, Patna | Approved | 06/11/2020 |
| IEC, Maharaja Agrasen Hospital | Approved | 25/12/2020 |
| Institutional Ethics Committee, King George Hospital, Visakhapatnam | Approved | 12/11/2020 |

| **Name of Committee** | **Approval Status** | **Date of Approval** |
| --- | --- | --- |
| Ethics Committee, St. Theresa’s Hospital | Approved | 07/06/2021 |
| Guru Teg Bahadur Hospital Ethics Committee | Approved | 03/06/2021 |
| IEC ESIC Medical College and Hospital, | Approved | 08/06/2021 |
| IEC Prakhar Hospital Pvt Ltd | Approved | 04/06/2021 |
| IEC, All India Institute of Medical Sciences, Patna | Approved | 09/06/2021 |
| IEC, Maharaja Agrasen Hospital | Approved | 06/06/2021 |
| Institutional Ethics Committee- AIG Hospital | Approved | 15/06/2021 |

6. **Institutional Ethics Committees (IECs) of 7 study sites for phase-2 (BECT069) study**

**References**.

1.	NIBSC. Data Sheet -Research reagent for anti-SARS-CoV-2 Ab NIBSC code 20/130.

2.	Bewley KR, Coombes NS, Gagnon L, et al. Quantification of SARS-CoV-2 neutralizing antibody by wild-type plaque reduction neutralization, microneutralization and pseudotyped virus neutralization assays. *Nat Protoc* 2021; **16**(6): 3114-40.

3.	Mattiuzzo G, Bentley EM, Hassall M, et al. Establishment of the WHO International Standard and Reference Panel for anti-SARS-CoV-2 antibody. *World Health Organization* 2020; **60**.

4.	Petrovsky N, Harrison LC. Cytokine-based human whole blood assay for the detection of antigen-reactive T cells. *J Immunol Methods* 1995; **186**(1): 37-46.
